# Quantitative microbial risk assessment of human H5N1 infection from consumption of fluid cow’s milk

**DOI:** 10.1101/2024.12.20.24319470

**Authors:** Katherine J. Koebel, Ece Bulut, Samuel D. Alcaine, Aljoša Trmčić, Mohammed Nooruzzaman, Lina M. Covaleda, Diego G. Diel, Renata Ivanek

## Abstract

The spillover of H5N1 clade 2.3.4.4b into dairy cattle has raised concerns over the safety of fluid milk. While no foodborne infection has been reported in humans, this strain has infected at least 70 people and milk from infected cows is known to be infectious by ingestion in multiple other species. Investigation into the public health threat of this outbreak is warranted. This farm-to-table quantitative microbial risk assessment (QMRA) uses stochastic models to assess the risk of human infection from consumption of raw and pasteurized fluid cow’s milk from the United States supply chains. These models were parameterized with literature emerging from this outbreak, then employed to estimate the H5N1 infection risk and evaluate multiple potential interventions aimed at reducing this risk. The median (5^th^, 95^th^ percentiles) probabilities of infection per 240-mL serving of pasteurized, farmstore-purchased raw, or retail-purchased raw milk were 7.66E-19 (2.39E-20, 4.02E-17), 1.56E-7 (6.67E-10, 1.28E-5), and 1.40E-7 (6.65E-10, 1.13E-05), respectively. Our results confirm that pasteurization is highly effective at reducing H5N1 infection risk. Scenario analysis revealed quantitative real-time reverse transcriptase-polymerase chain reaction (qrRT-PCR) testing of bulk tank milk to be an effective method for numerically reducing risk from raw milk. Additionally, we identify knowledge gaps related to human H5N1 dose-response by ingestion and raw milk consumption patterns. These findings emphasize the importance of mechanistic epidemiologic models for informing public health responses amidst outbreaks with foodborne potential and highlight the need for additional research into raw milk consumption patterns to better understand this exposure pathway.

## 1. INTRODUCTION

Avian influenza (AI) represents a serious risk to both avian and mammalian species. AI is an illness caused by multiple strains of the influenza A virus (IAV) (Alexandar, 2007; Swayne, 2023). Of particular interest are the viruses of the H5N1 clade 2.3.4.4b, which emerged in 2020 and were subsequently detected in North America in late 2021 (Bevins et al., 2021; Kaneil et al., 2023). Epidemiologic data highlight the predominance of this clade in reports of influenza A(H5) samples from both humans and animals (WHO, 2024; WHO, 2023).

In March 2024, a spillover event resulted in the infection of dairy cattle in Texas with H5N1 clade 2.3.4.4b (Caserta et al., 2024; Burrough et al., 2024). The initial infections have since spread to more than 1,000 herds nationwide as of 26 September 2025 (USDA-APHIS, 2025b). Infected cows present predominantly with clinical mastitis, usually in mid-to late-lactation, with additional signs such as anorexia, rumen hypomotility, and nasal discharge (Caserta et al., 2024; Burrough et al., 2024). Milk from these animals contains extremely high concentrations of live virus (4.0-8.8 log_10_TCID_50_/mL (tissue culture infectious dose 50, hereafter referred to as “logTCID_50_”).

As of 26 September 2025, out of 70 total human cases, 41 were attributed to dairy cattle exposure in addition to 1 mortality with “other animal exposure” (Uyeki et al., 2024; CDC, 2025a). Human cases are generally mild, including conjunctivitis and upper respiratory symptoms, and no human-to-human transmission has been reported (Neumann et al., 2024).

Exposure in 2 cases is unknown (CDC, 2025a). No confirmed cases have been definitively attributed to consumption of dairy products, although two raw milk operations issued product recalls after their cattle were determined to be infected. The resultant risk to consumers was acknowledged by public health agencies (LA Public Health, 2024; CDPH, 2024).

While primarily considered respiratory pathogens, evidence exists that avian influenza viruses (AIVs), including H5N1, are infectious via the gastrointestinal (GI) tract, and that human infection via the foodborne route is plausible. Bullock et al. (2025) provide a comprehensive review of this evidence, highlighting successful experimental H5N1 infections in cats, swine, guinea pigs, hamsters, mice, and ferrets after oral, orogastric, or intragastric inoculation.

Previous research has also proven the ability of H5N1 to bind to sialic acid receptors in human GI tissue (Shu et al., 2010) and demonstrated viral replication in the intestines of a pediatric patient that died of H5N1 (Uiprasertkul et al., 2005). Milk from cows infected with H5N1 clade 2.3.4.4b is known to be infectious by ingestion in mice (Eisfeld et al., 2024; Guan et al., 2024) and by orogastric inoculation of cynomolgus macaques (*Macaca fascicularis*) with a bovine isolate establishing subclinical infection with seroconversion (Rosenke et al., 2025). It is thought that exposure of the pharynx to, and/or absorption via GI lymphatics of, infectious virions contributes to the establishment of infection (Bullock et al., 2025; Rosenke et al., 2025; Shinya et al., 2011). Human cases with known cattle exposure cite contact with raw milk prior to illness (Morse et al., 2024). The epidemiological study by Garg et al. (2024) of the Centers for Disease Control and Prevention (CDC) additionally found that, among 25 dairy workers who contracted avian influenza, “4 (16%) were exposed to cows and 21 (84%) to both cows and raw milk;” in their Table 1 they additionally clarify that “raw milk” refers to raw milk consumption, raw milk exposure, or both.

**TABLE 1.**
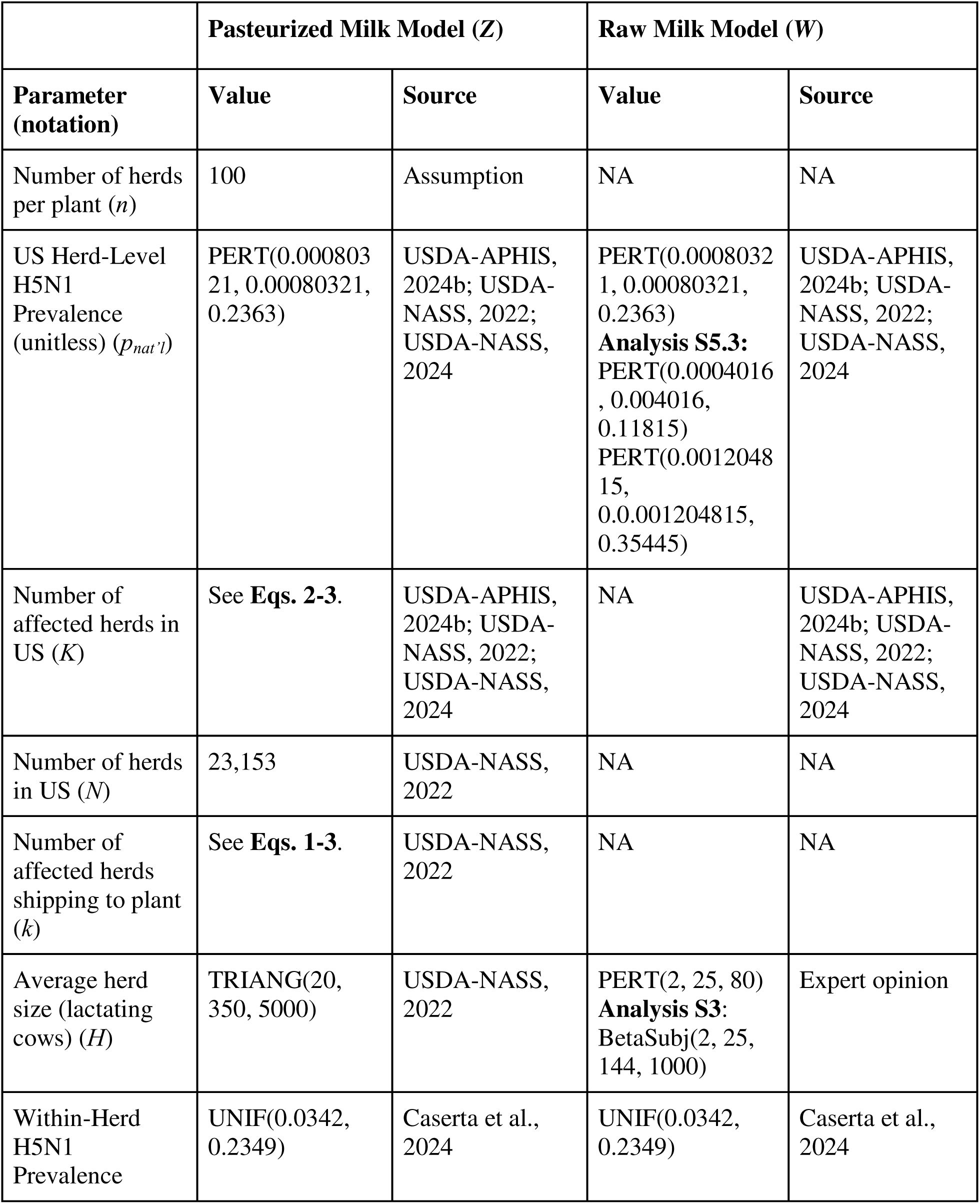
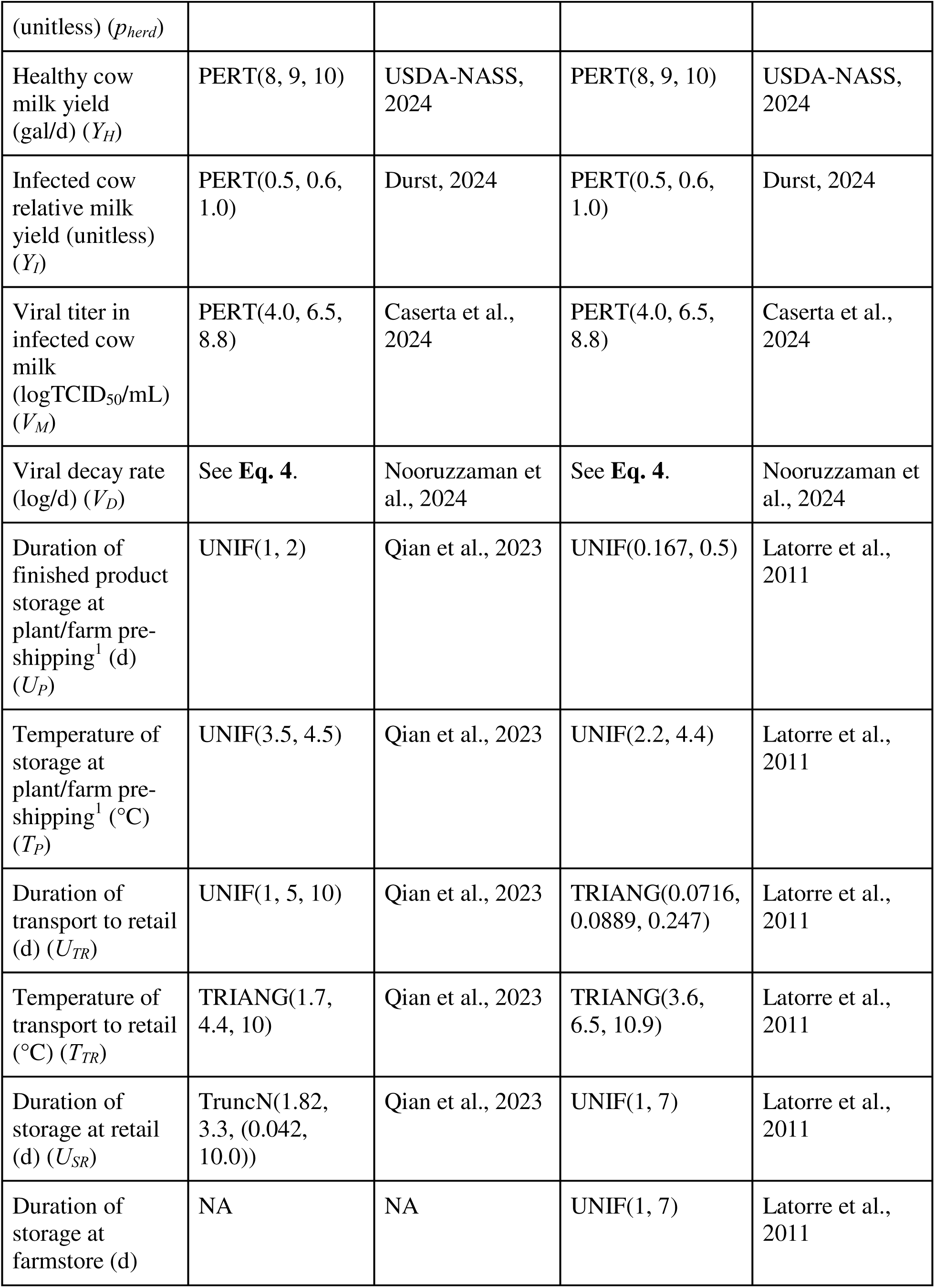
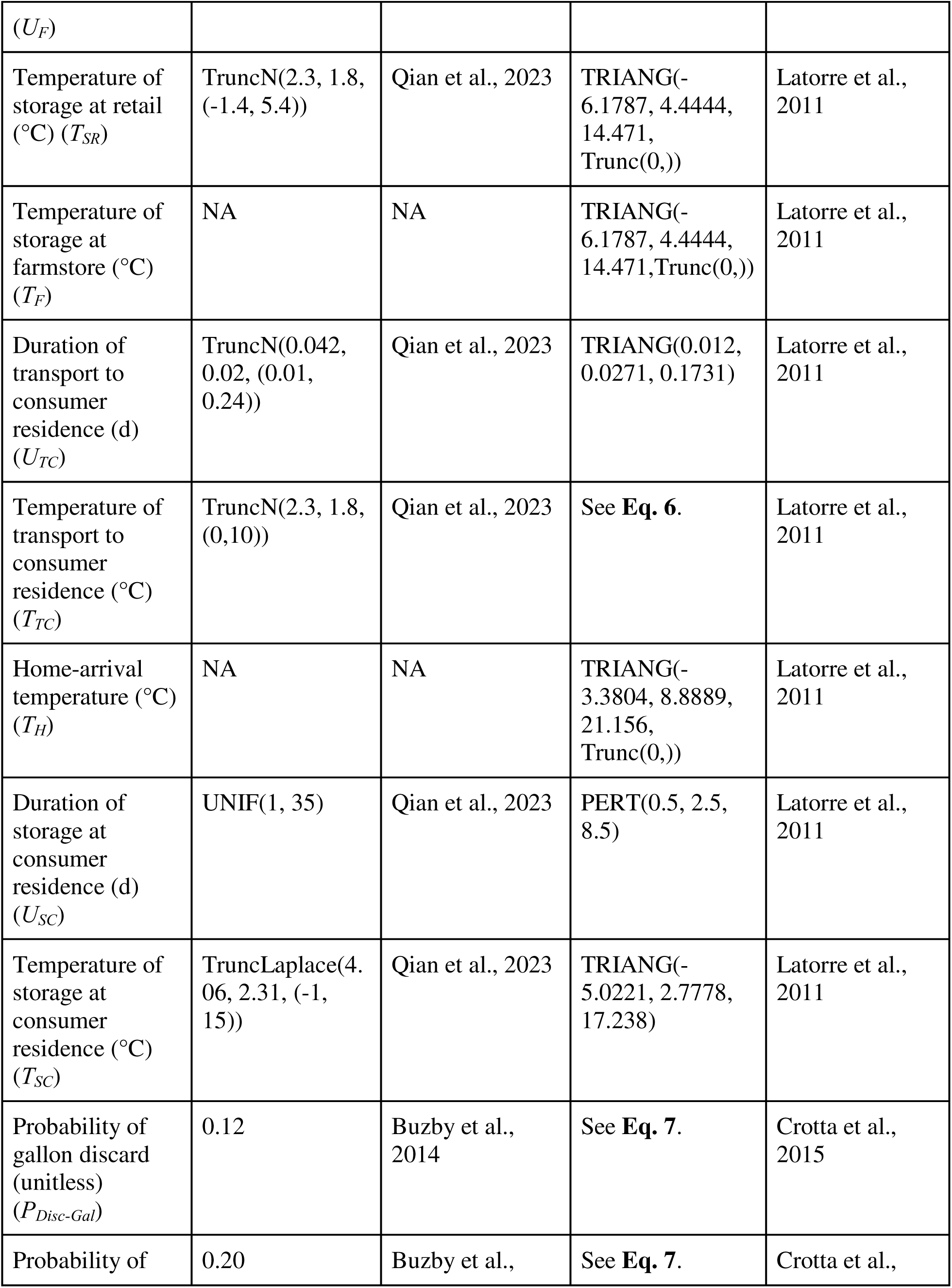
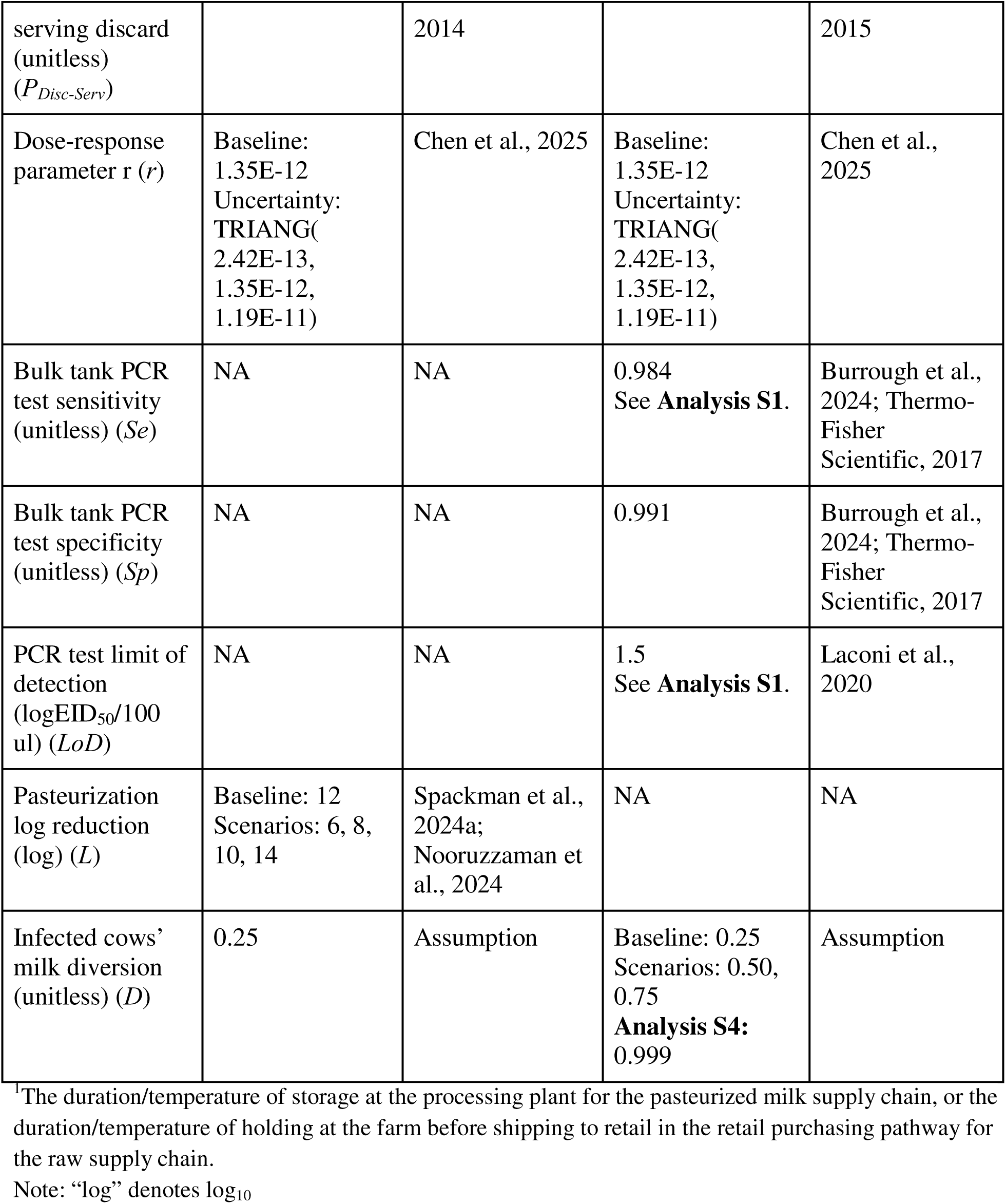
Parameters used in a quantitative risk assessment of human H5N1 infection from consumption of pasteurized or raw fluid cow’s milk.

Given the plausibility of human infection by ingestion and the demonstration of copious shedding from infected cows, questions remain about the risk of human H5N1 infection from consuming fluid dairy milk. Heat inactivation assays in milk demonstrate efficacy (Guan et al., 2024; Nooruzzaman et al, 2025a); for example, high-temperature short-time (HTST) pasteurization is posited to yield a minimum 12-log reduction in viral loads (Spackman et al., 2024a). Despite the effectiveness of pasteurization in reducing the risk of foodborne illness, consumption of raw milk remains popular in the United States (US); 4.4% of US adults consume raw milk at least once per year (Lando et al., 2022; CDC, 2024b; FDA, 2024b).

Research into the human H5N1 infection risk associated with fluid milk consumption is critical to inform efforts aimed at reducing said risk in the current and potential future outbreaks, including updates to public health policy or implementation of surveillance strategies. Existing risk assessments report the risk to pasteurized milk consumers as negligible to low, and moderately uncertain, while the risk to raw milk consumers is numerically higher (Chen et al., 2025; Browne et al., 2025). However, these publications are either qualitative in nature and emphasize international rather than domestic populations or fail to represent farm-to-table dynamics that may mechanistically impact risk, such as diversion of infected cows or viral decay during storage. Thus, the objectives of this study were to (i) estimate the risk of human H5N1 infection from consumption of raw or pasteurized fluid milk during the early epidemic (March-October 2024), (ii) assess the impact of multiple potential interventions on baseline risk, and (iii) identify and scrutinize knowledge gaps related to milkborne human infection risk.

## 2. METHODS

To evaluate the extent of the risk posed to milk consumers by the bovine H5N1 outbreak, we performed a quantitative microbial risk assessment (QMRA). In accordance with the Codex Alimentarius (FAO, WHO, 2014), a QMRA consists of four systematic steps: (i) hazard identification, (ii) hazard characterization, (iii) exposure assessment, and (iv) risk characterization. These guidelines formed the basis of our analytical approach, with additional considerations from Boles et al. (2022) and ACMSF (2012). The purpose of this QMRA was to estimate the risk of human H5N1 infection per individual 240 mL (8 fluid ounce) serving of fluid dairy milk of unspecified fat percentage. This involved construction of two bottom-up stochastic QMRA models (Bosch et al., 2018), leveraging the structures of previously described pasteurized (Qian et al., 2023) and raw milk (Latorre et al., 2011) supply chains. The models were built in Microsoft Excel 365 v.2406 (Microsoft Corp., Redmond, WA) with @RISK v8.6.1 (Lumivero, Denver, CO). The main outcome of interest was the probability of infection per serving (hereafter “p(infection)”): a function of the probability of serving contamination (“p(serving)”) and the quantity of virus in said serving (“Q_serv_”). We utilize median risk accompanied by 95% confidence intervals (CI) as the primary metric (Hunter et al., 2010). We also report the projected annual number of milkborne H5N1 infections, as detailed in *2.4 Risk Characterization*.

### 2.1 Hazard Identification

In a study of n=297 pasteurized retail dairy product samples from 17 states, viral RNA from bovine H5N1 was detected via PCR in approximately 20% of fluid milk samples, but no infectious virus was detected by egg inoculation (Spackman et al., 2024). However, the live virions present in raw milk from infected cows are known to be infectious *in vitro* (Guan et al., 2024) and *in vivo* (Guan et al., 2024; Eisfeld et al., 2024). The strain of H5N1 responsible for the outbreak is infectious in humans (Morse et al., 2024; Uyeki et al., 2024; CDC, 2025a), and exposure to raw milk is a reported risk factor (Garg et al., 2025). Finally, research in both animal models and human cells supports the hypothesis that H5N1 is infectious via GI exposure (Rosenke et al., 2025; Shu et al., 2010; Bullock et al., 2025; Eisfeld et al., 2024; Guan et al., 2024, Shinya et al., 2011; Lipatov et al., 2009). As such, there is sufficient evidence to warrant investigation into the risk of human H5N1 infection via fluid dairy milk products due to the 2024 spillover event. Our position aligns with the current US government’s investment in preventing consumer exposure through control strategies, including a Federal Order in April 2024 requiring cattle be tested prior to interstate movement, and (ii) a December 2024 Federal Order that implemented mandatory testing/reporting under the National Milk Testing Strategy (USDA, 2024). The latter Federal Order released by USDA-APHIS (2024) states verbatim that “APHIS has determined that good cause exists to implement these additional requirements without notice and comment, as evidence has shown that raw, unpasteurized milk is a vehicle for the spread of the HPAI H5N1 virus.” They also emphasize that, “while studies have confirmed pasteurized milk and dairy products are safe, raw, unpasteurized milk consumption remains a risk for multiple diseases including HPAI.”

### 2.2 Exposure Assessment

Two separate models were developed to represent the US pasteurized and raw fluid milk supply chains from farm to table. This was done due to fundamental differences in these supply chains, including variations in processing (i.e. pooling of milk from multiple farms), differences in the constituent steps of the cold chain, and distinct distributions for multiple parameters (e.g. herd size). In the raw milk model, two purchasing pathways (farmstore and retail) were differentiated to represent the two available avenues for obtaining raw milk in the US as these pathways also differ in their cold chain parameters (Latorre et al., 2011). However, both models function similarly with 3 main components: Herd Dynamics, Processing & Packaging, and Consumption & Infection. See **Figure 1** for a conceptual model and **Table 1** for a complete list of parameters.

**FIGURE 1.**
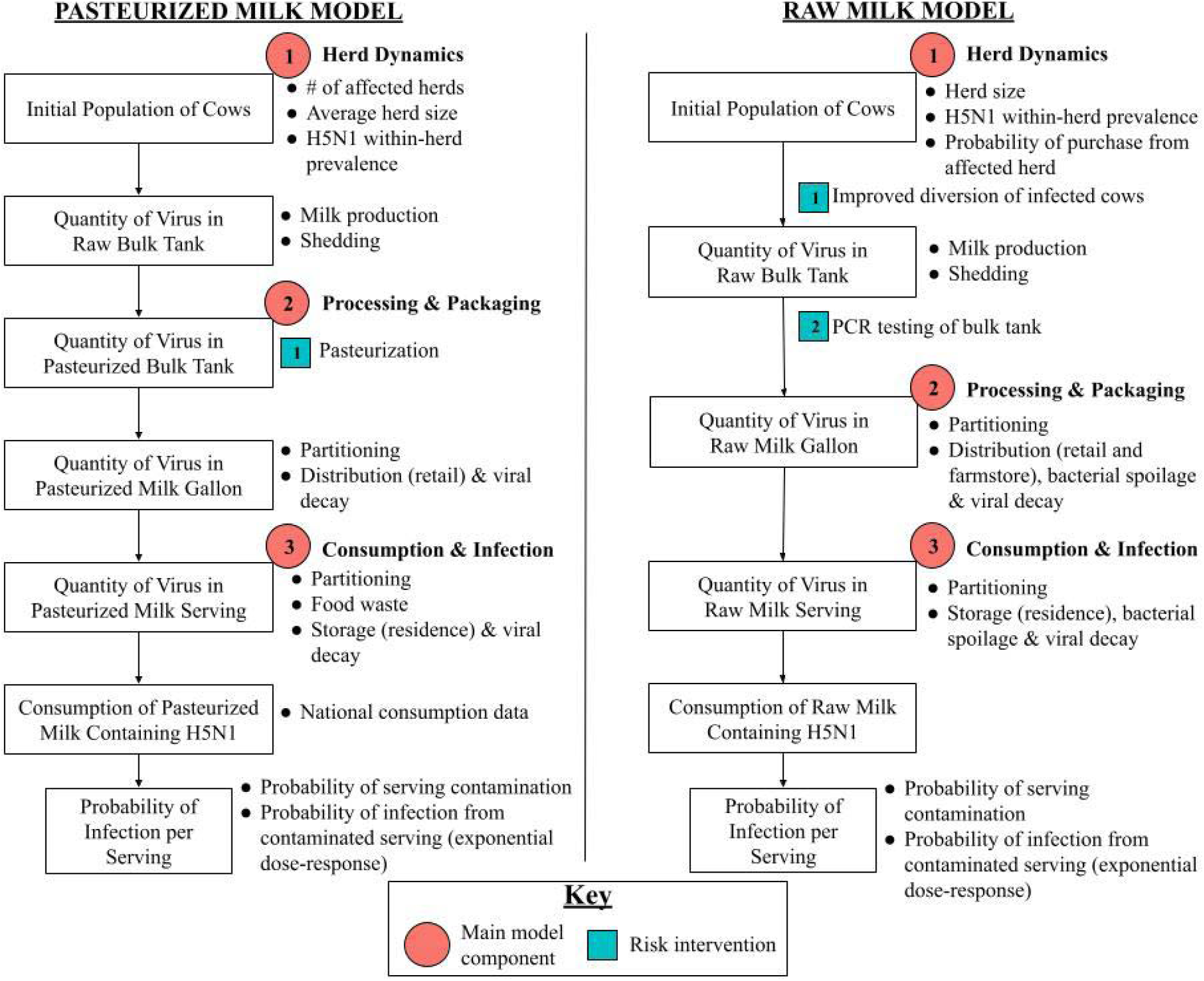
Conceptual model for a quantitative risk assessment of human H5N1 infection from consumption of pasteurized or raw fluid cow’s milk.

#### 2.2.1 Pasteurized Milk Model

This model represents a given fluid milk processing plant receiving, processing, packaging, and distributing milk from a fixed number of herds. Fluid milk is followed from its harvest at the contributing farms, through processing at the facility, distribution through retail, and culminates in its consumption by a consumer as an individual serving.

The lactating cow herds are of average size *H_Z_* with a healthy cow producing *Y_H_* gal/d (USDA-NASS, 2022, 2024). Infected cows comprise *p_herd_* proportion of the herd and give *Y_I_*relative yield (%) with milk titers *V_M_* (logTCID_50_/mL) (Caserta et al., 2024; Durst, 2024). A proportion *D* (%) of these animals is identified and their milk is discarded at the time of harvest (i.e. does not enter the bulk tank). Diversion of this milk is understood to mean identification of the animal as unfit to contribute to the milk supply, including but not limited to recognition of clinical illness or laboratory confirmation of H5N1 infection.

The plant receives milk from 100 herds (*n*), where the number of affected herds (*k*) shipping to that plant is calculated with a hypergeometric distribution such that:

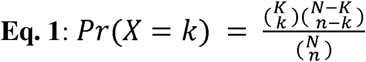

where *N* is the number of dairy herds in the US (23,153) (USDA-NASS, 2022) and *K*, the hypergeometric distribution parameter for the number of successes (i.e. affected herds) in the population, is represented by:

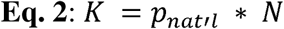

Using APHIS detection data obtained on November 5, 2024 (USDA-APHIS, 2024b) and the USDA 2022 Census of Agriculture (USDA-NASS), the US herd-level H5N1 prevalence parameter *p_nat’l_* was calculated from values of the monthly period prevalences for states (*p(state.month)*) with confirmed H5N1 infections in dairy herds such that:

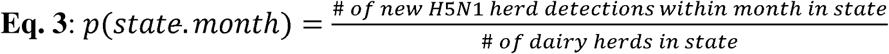

by obtaining the number of herds detected positive in a state from the aforementioned APHIS detection data (USDA-APHIS, 2024b), and the total number of herds in that state from the USDA 2022 Census of Agriculture (USDA-NASS). Next, a PERT distribution was fitted with the minimum, most likely and maximum values of *p(state.month)* to form *p_nat’l_*. In this approach, the state-month prevalence values used in parameterization of *K* (and the US herd-level H5N1 prevalence, *p_nat’l_*) indirectly account for the clustering phenomenon of localized outbreaks amongst herds in geographic proximity supplying milk to the same processing plant (**Assumption 2**).

The probability that a processing plant receives milk from at least one infected herd is modeled with **Eq. 1** as Pr(*X* > 0) in each iteration. Milk is pooled at the plant and held 24 hours, during which the viral titers undergo logarithmic decay *V_D_*. Temperature-dependent daily log decay (logTCID_50_/d) is calculated such that:

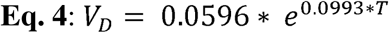

where *T* is the temperature parameter (°C) and *V_D_*is the log (base-10) decay rate (logTCID_50_/d) during the corresponding cold chain step. This model was fit using empirical data from Nooruzzaman et al. (2025a), which consisted of viral decay rates in replicates run in fluid milk at 4, 20, 30, and 37°C (**Analysis 5** in *Supplementary Information*). **Eq. 4** was applied to all storage/transportation stages in both parameterized and raw models.

The milk is HTST pasteurized, which is estimated to yield >12-log reduction of H5N1 (Spackman et al., 2024a). Milk is packaged into gallon (3,785 mL) containers and the partitioning of viruses, including the resultant quantity in contaminated containers and the probability of container contamination, is modeled according to the binomial distribution and gamma function, respectively (Nauta 2005). As we assume that virions do not cluster in fluid milk (**Assumption 3**).

Gallons proceed through the cold chain: storage at the plant (*U_P-Z_, T_P-Z_*), transport to retail (*U_TR-Z_, T_TR-Z_*), storage at retail (*U_SR-Z_, T_SR-Z_*), transport to the consumer residence (*U_TC-Z_, T_TC-Z_*), and storage at the residence (*U_SC-Z_,* and *T_SC-Z_*) (Qian et al., 2023). Duration parameters are expressed in days (d) and temperature parameters in °C (**Table 1**). Food waste of gallons (*P_Disc-Gal-Z_*) and servings (*P_Disc-Serv-Z_*) is accounted for (Buzby et al, 2014).

#### 2.2.2 Raw Milk Model

As aforementioned, all 3 model components (Herd Dynamics, Processing & Packaging, and Consumption & Infection) are shared with the pasteurized milk model, with several adaptations. Specifically, the raw milk model represents a single given herd of size *H_W_* (lactating cows) selling raw fluid milk through either farmstore or retail pathways modeled in parallel. The parameter *H_W_* was parameterized as PERT(2,25,80), based on the input from two dairy practitioners with more than 10 years of experience serving farms that sell raw milk in NY, who provided their expert opinions that the vast majority of farms selling raw milk for human consumption in the US are very small operations, ranging between 2 and 80 cows, with an average of 25 cows. No pooling of milk from multiple herds nor pasteurization occurs. The probability that the herd is infected is given by parameter *p_nat’l_* (**Eq. 3**). Sharing parameter *p_nat’l_* between the pasteurized and raw milk models is driven by the assumption that raw milk herds do not systematically differ from the national average (**Assumption 4**).

Parameters *p_herd_*, *Y_H_, Y_I_, V_M_, V_D_, D,* and *r* are shared with the pasteurized milk model. In the farmstore pathway, gallon packages are sold directly to consumers after storage at the farmstore (*U_F_, T_F_*). In the retail pathway, on-farm storage lag (*U_P-W_, T_P-w_*), retail transport (*U_TR-W_, T_TR-W_*), and retail storage (*U_SR_*, *T_TR_*) are modeled (Latorre et al., 2011). In both pathways, packages are transported to (*U_TC-W_, T_TC-W_, T_H_,*) and stored at (*U_SC-W_, and T_SC-W_*) the consumer residence. Temperature parameters remain expressed in °C and duration parameters in days (d) as in the pasteurized milk model. The milk temperature during transport to the residence (*T_TC-W_*) is calculated with:

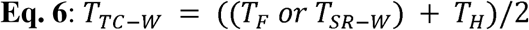

where *T_F_* and *T_SR-W_* are the temperatures of storage at the farmstore or retail, respectively, *T_H_* is the temperature of home arrival, and *T_TC-W_* is the temperature during transport to the consumer residence (°C) (Latorre et al., 2011).

Accounting for raw milk perishability, the probability of spoilage and discard of the product during a given step in the cold chain is adapted from Crotta et al. (2015) and modeled using the general formula:

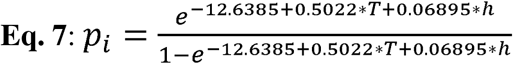

where *T* is the temperature parameter (°C), *h* is the duration parameter (d), and *p_i_*is the probability of spoilage and subsequent discard of a gallon (*P_Disc-Gal-W_*) or serving (*P_Disc-Serv-W_*).

A 240 mL serving is partitioned from the package and immediately drank by a consumer. The probability that a consumed serving was contaminated (p(serving)) and the quantity of virus in a contaminated serving (Q_serv_ (TCID_50_)) are calculated with the aforementioned mathematical partitioning methodology of Nauta (2005).

#### 2.2.3 Validation of the exposure assessment and model assumptions

The QMRA predictions must be interpreted in the context of the models’ validation and assumptions. Validation of the exposure assessment in the pasteurized milk model was performed using the surveillance data of Spackman et al (2024b). They report the concentration of inactivated viral material in EID_50_/mL in whole (3.0±1.1), 2% (3.1±1.2), 1% (3.1±1.2), and skim milk (3.3±0.7) after backcalculation via the results of quantitative real-time reverse transcription polymerase chain reaction (qrRT-PCR) testing: a molecular diagnostic method based on the presence of viral RNA. Nooruzzaman et al. (2025a) demonstrated that heat treatment does not affect the quantity of viral RNA remaining in the product, providing support for our assumption that viral material (i.e. inactivated virus) in the presence of pasteurization approximates the number of viable viruses in the same milk in the absence of pasteurization (and vice versa). Moreover, paired t-testing of EID_50_ and TCID_50_ titers for H5N1 clade 2.3.4.4b in milk by co-authors MN and DD revealed no significant difference between these values (p=0.15), supporting that these metrics can be considered approximations of each other (see **Analysis S2** in *Supplementary Materials*). The geographic origin of milk samples analyzed in Spackman et al. (2024b) is unknown. Thus, using the *homogeneous* herd distribution approach (**Assumption 2**) we set *K*=26, representing the number of H5N1-affected herds in the US during the month (i.e. April 2024) in which their surveillance was conducted. Parameter *L* was set at 0 so that the model would not “remove” any viruses during the pasteurization step, allowing us to predict the corresponding concentration of inactivated virus in pasteurized milk (see **Figure S3**).

Therefore, the pasteurized milk model output reporting the concentration of inactivated virus (TCID_50_/mL) in milk at retail could be interpreted as the equivalent amount of inactivated virus (i.e., viral genetic material remaining in the product post-pasteurization) in the Spackman et al. (2024b) study, thus approximating their reported metric. The outputs for this value in this simulation were compared graphically against the empirical data (**Figure 2**). Note that as our model does not differentiate between fat percentages in fluid milk and that the results from all fluid milk varieties (e.g. skim, 1%, 2%, and whole) reported in the empirical data were considered together in this comparison.

**FIGURE 2.**
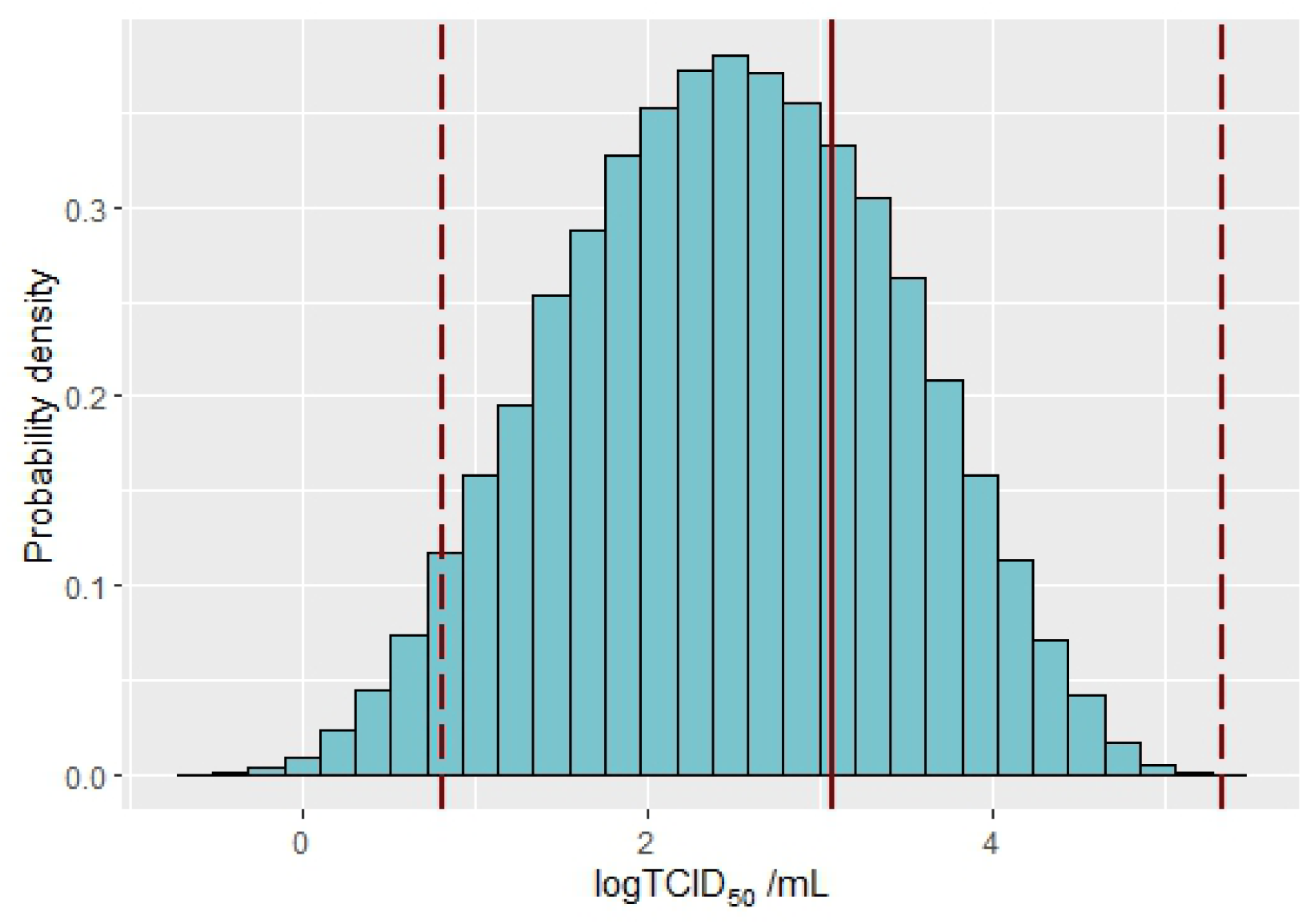
Estimated concentration of inactivated H5N1 virus in milk gallon containers at retail in the pasteurized supply chain. The probability density histogram shows the model prediction in log_10_TCID_50_/mL. Overlaid vertically are the mean (solid line) concentration of H5N1 virus ± 2 standard deviations (dashed lines) reported in logEID_50_/mL by Spackman et al. (2024b), which were arrived at after backcalculation from PCR testing. As pasteurization does not remove viral RNA, qrRT-PCR performed on pasteurized products serves as a proxy for the concentration of virus in the product, albeit inactivated. We approximated this metric by running our model with pasteurization parameter *L*=0, allowing us to predict the concentration of (inactivated) virus in milk gallons, thus mimicking the backcalculation approach in Spackman et al. This approach assumes log_10_TCID_50_/mL and logEID_50_/mL approximate each other as supported by **Analysis S2**.

We identified 4 assumptions meriting further analysis. The fundamental assumption underlying this work is **Assumption 1**: that milk from H5N1-infected cows is infectious to humans via ingestion through an exponential dose-response mode (see **Eq. 5** below).

**Assumption 1** is scrutinized with citation of relevant literature in *Discussion* and an uncertainty analysis of the dose-response parameter *r*. **Assumption 2** was analyzed by replacing the calculation of parameter *K* with a distribution directly from historical herd incidence data, thereby modeling geospatial homogeneity. We tested **Assumption 3** by manipulating the virion clustering parameter. Finally, we evaluated **Assumption 4** by shifting the probability of herd infection for raw milk herds to be systematically lower or higher than that for conventional herds. The resultant values for p(infection) were compared numerically and graphically against the baseline. In the interest of space, additional details regarding the assumption analysis methodologies are included in *Supplementary Materials*.

### 2.3 Hazard Characterization

In another milkborne H5N1 infection risk assessment, authors adapt an exponential model for the inhalational human influenza A dose-response curve and translate parameter *r* upward to represent viral reduction from acidic conditions in the stomach (Chen et al., 2025). Therein, the authors adapt an existing inhalational exponential dose-response model for H5N1 in humans and its associated uncertainty range for parameter *r*, and pair this with the results of a literature review examining the impact of gastric conditions (e.g., pH, emptying) on IAV viability. Based on their findings, the authors deemed a 3-log translation appropriate to shift parameter *r* (1.35E-12) and its uncertainty range (2.42E-13 to 1.19E-11) to represent viral inactivation during gastric passage. In the current study, we adopt the Chen et al. approach and use an exponential dose-response model, defined as:

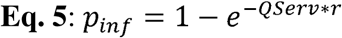

where *Q_Serv_*is the count of TCID_50_ present in a contaminated serving as predicted in the exposure assessment, *r* is the dose-response parameter, and *p_inf_* is the resultant probability of infection from the contaminated serving. To separate uncertainty from variability, we set *r* at 1.35E-12 in the baseline scenario and performed a separate analysis with a simulation in which *r* was allowed to vary within TRIANG(2.42E-13, 1.35E-12, 1.19E-11) alongside all other stochastic model parameters.

### 2.4 Risk Characterization

The probability of infection (p(infection)) from consuming any serving was calculated as a product of the probability that the consumed serving was contaminated, p(serving), and the probability of developing infection as a result of the virus quantity in said contaminated serving, *p_inf_*. Thus, p(infection) may be interpreted as the probability of H5N1 infection from any given serving of fluid cow’s milk.

We calculated the projected annual number of milkborne human H5N1 infections by multiplying p(infection) in the pasteurized and raw milk models by the respective number of 240 mL pasteurized and raw fluid milk servings consumed in the US per year. Consumption statistics were obtained from the QMRA by Chen et al. (2025); the authors report the daily number of “average” (212 mL) pasteurized and raw fluid milk servings consumed in the US. We adapted this number to our QMRA by adjusting for a standardized 240 mL serving size by calculating their total daily volume consumption and dividing this by 240, then multiplying by 365 to obtain the total number of 240 mL pasteurized and raw fluid milk servings consumed annually in the US.

Each simulation consists of 50,000 iterations generated with Latin hypercube sampling and Mersenne twister pseudorandom number generation. Convergence testing with 5% tolerance and 95% confidence confirmed the sufficiency of the iteration number. Figures, tables, and analyses were produced in R v4.3.2 (R Foundation for Statistical Computing, Vienna, Austria).

To identify model parameters most influential on p(infection) representing knowledge gaps or potential intervention points for public health policy, we conducted a sensitivity analysis and report the partial rank correlation coefficients ( ) achieving statistical significance (p < 0.05/(number of stochastic parameters in model); 16 in the pasteurized milk model and 12 for both purchasing pathways for the raw milk model) for the baseline scenarios for pasteurized and raw milk. Parameters with | | ≤ 0.1 were considered negligible with low practical significance (Schober et al., 2018) and were omitted from further discussion. Interpretation of the coefficient strength ranges was adapted from this source as well, wherein 0.1 < | | < 0.40 is weak, 0.40 ≤ | | < 0.70 is moderate, and 0.70 ≤ | | is strong.

We performed scenario analyses to determine the impact of, in the pasteurized milk model, (i) pasteurization efficacy, and in the raw milk model, (ii) bulk tank PCR testing and (iii) improved preharvest diversion of infected cows on p(infection). The purpose of this approach was to evaluate the risk mitigation effects of pasteurization, bulk tank PCR testing, and improved animal-level screening in reducing milkborne H5N1 infection risk.

In the pasteurized milk model, the pasteurization log reduction parameter *L* was tested at levels 6-, 8-, 10-, 12-(baseline), and 14-log reduction. A minimum level of 6 was chosen based on the thermal inactivation assays of Nooruzzaman et al. (2025a), in which HTST pasteurization conditions were concretely demonstrated to completely inactive H5N1 clade 2.3.4.4b in raw clinical milk samples of approximately 6 logTCID_50_/mL starting concentration.

A full factorial scenario analysis was performed to examine two interventions aimed at reducing risk from consumption of raw milk. Diversion parameter *D* was evaluated at baseline 25%, and levels 50% and 75%, reflecting an unspecified “improved ability” to remove the milk of infected cows from the supply chain before harvest. These levels were tested in combination with implementation of once-daily PCR testing of bulk tank milk. Parameters sensitivity (*Se*), specificity (*Sp*), and limit of detection (*LoD*) are employed regarding PCR testing (Thermo-Fisher Scientific, 2017; Laconi et al., 2020). The model simulates the once-daily collection of a single 0.10 mL aliquot directly from the bulk tank for laboratory testing. Partitioning of spatially uniformly distributed viruses into this quantity utilizes the same methodology for partitioning bulk tank milk into gallon containers and from gallon containers into an individual serving. The bulk tank milk is held for 24h while the aliquot is transported to the laboratory and tested, with viral decay *V_D_* during this lag time as previously mentioned (Nooruzzaman et al., 2025a). Bulk tanks testing true (at a probability *Se*) or false (at a probability 1-*Sp*) positive by PCR are dumped and contribute no risk. Tanks testing negative either by false negative (at a probability 1-*Se*) or falling under the *LoD* proceed through the remainder of the model. An additional, truncated analysis into the impact of a test with improved analytic and/or diagnostic sensitivity is provided in **Analysis S1**. **Analyses S3** and **S4** examine the effect of larger raw milk herd sizes and relative impact of bulk tank PCR testing versus improved cow diversion, respectively.

Issues surrounding dose-response assessment remain leading causes of concern for risk analysts (Hamilton et al., 2024). The dose-response mechanism in our models is similarly a source of uncertainty and, as such, merited further scrutiny. As explained in **2.3 Hazard Characterization**, we subjected the dose-response parameter *r* to an uncertainty analysis.

## 3. RESULTS

### 3.1. Validation of Pasteurized Model

Projections for the concentration of inactivated virus (Q_serv_; (TCID50)) in gallon packages of pasteurized fluid milk at retail are shown to align with the empirical data from previously conducted surveillance testing (Spackman et al., 2024b). By examination of **Figure 2**, it is evident that the distribution for the pasteurized milk model’s prediction fall well within the Spackman et al. (2024b) reported range for titers as given by the mean ± 2 standard deviations.

As such, the pasteurized fluid milk model (its exposure assessment component) was considered validated to the extent possible given available data.

### 3.2. Baseline Scenario

The predicted risk metrics in the pasteurized milk model were very low (**Figure 3**). The 5th and 95th percentiles for p(infection) were 2.39E-20 and 4.02E-17, respectively, with a median value of 7.66E-19 and mean of 8.71E-18 (**Table S1**). The mean risk is 1 milkborne infection per 114,800 trillion servings. The quantity of virus per contaminated serving was similarly low, ranging from 0.00 to 0.30 logTCID_50_ with a mean value of 0.02 logTCID_50_. A value of 0.00 logTCID_50_ corresponds to 1 TCID_50_ in a serving and 0.30 logTCID_50_ corresponds to 2 TCID_50_ in a serving. Probability of serving contamination took percentiles of 2.43E-8 (5th) and 4.03E-5 (95th) with a median of 7.67E-7 and a mean of 8.66E-6 corresponding to 1 contaminated serving per 115,471. Note that this refers to the presence of live, infectious viruses and not viral RNA alone. When these predictions for the risk per any serving were scaled up to predict the number of human infections annually from consumption of pasteurized milk in the US, all iterations predicted zero infections.

**FIGURE 3.**
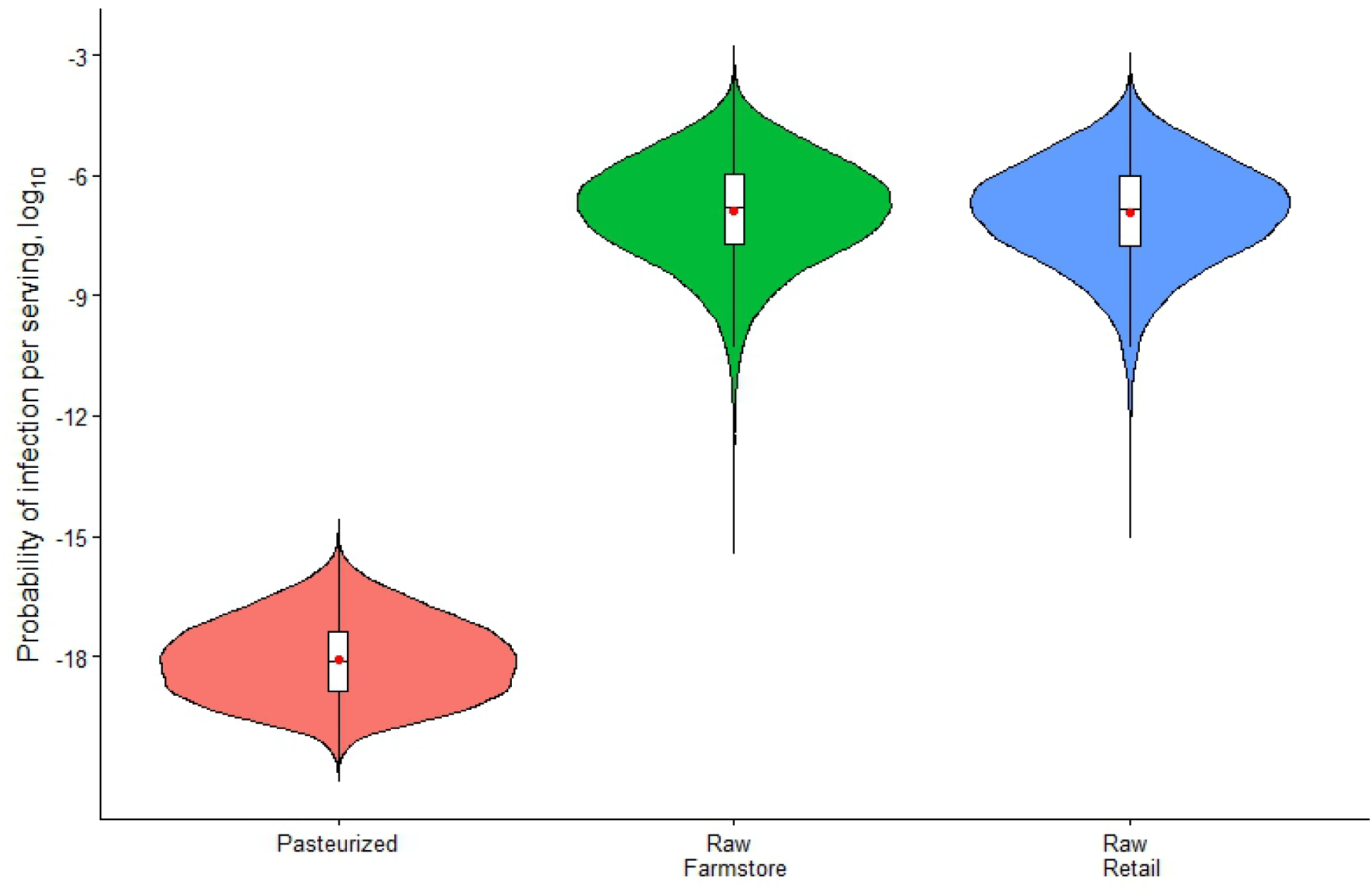
Violin plots for the baseline probability of H5N1 infection per serving (p(infection)) of pasteurized or farmstore- or retail-purchased raw milk. For the purposes of visualization, a logarithmic (base-10) Y-axis is employed. The mean is denoted with the red dot.

**FIGURES 4A-C.**
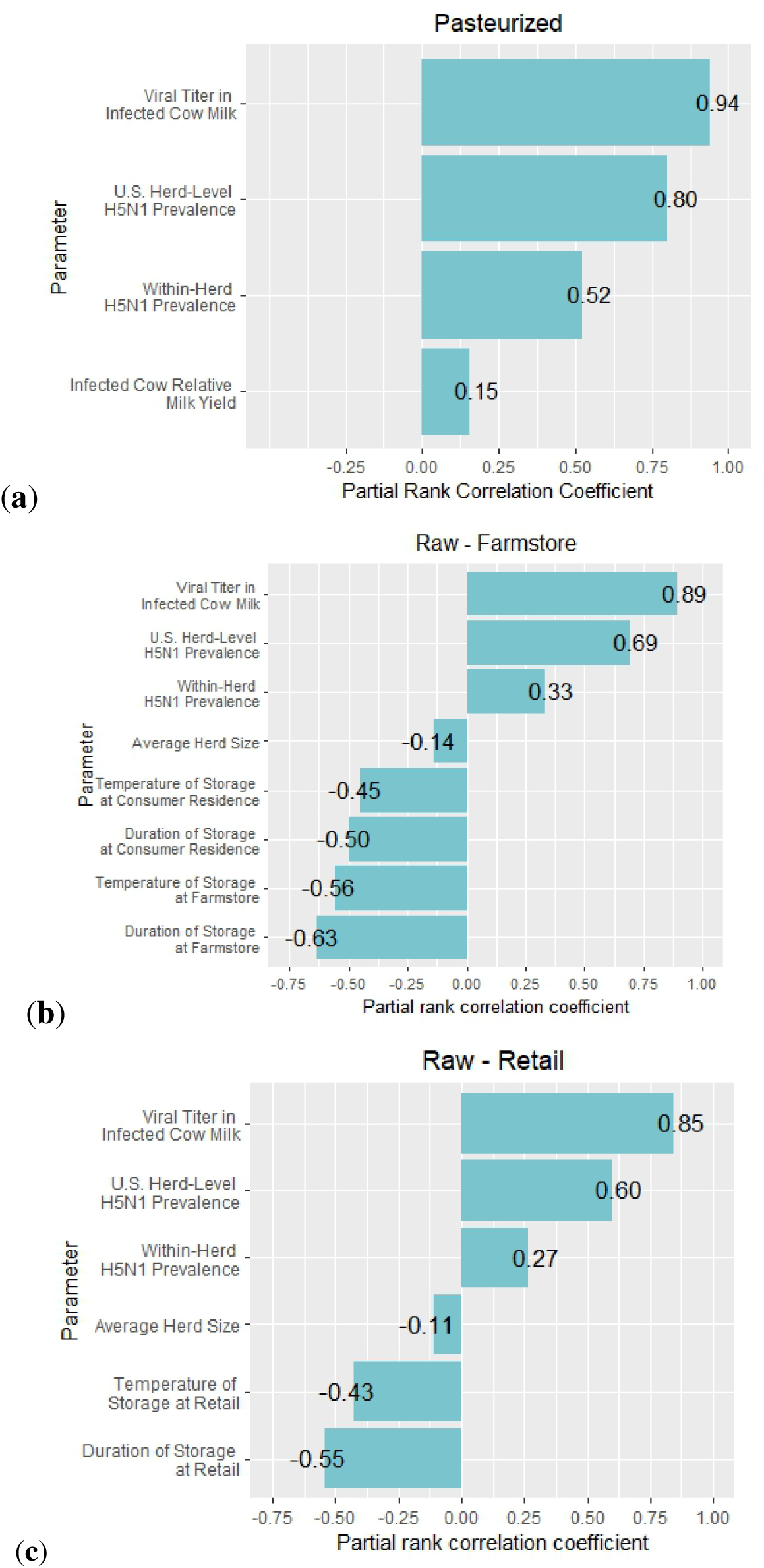
Sensitivity of the probability of infection per serving (p(infection)) of (a) pasteurized or raw milk obtained through (b) farmstore or (c) retail sale to model parameters achieving statistical (Bonferroni-corrected p ≤ 0.05/number of evaluated parameters in the model) and practical (correlation coefficient |⍰| > 0.1) significance.

In the raw milk model, risk metrics were considerably higher (12-log difference in medians) than in the pasteurized milk model (**Figure 3** and **Table S1**). Median p(infection) for the farmstore purchase pathway was 1.56E-7, with percentiles of 6.67E-10 (5th) to 1.28E-5 (95th) and a mean of 2.99E-6. Percentiles for the quantity of virus per contaminated serving ranged from 5.19 (5th) to 8.47 (95th) with a median value of 6.86 and mean 6.85 logTCID_50_. Median probability of serving contamination was 3.13E-2 (5th percentile: 3.21E-3; 95th percentile: 1.07E-1; mean: 4.00E-2 or, 1 contamination per 25 servings). Baseline p(infection) indicated negligible differences in risk between the two raw milk purchasing pathways (**Figure 3** and **Table S1**), likely due to similarities in the duration and temperature of the cold chains that reduce risk through probability of spoilage and subsequent discard. Paired with the increased access to farmstore-sold over retail raw milk in terms of state legality (Rhodes et al., 2019), further discussion of results is limited to the farmstore pathway, except in sensitivity analysis. For farmstore-purchased raw milk, the median predicted risk of infection per serving can be interpreted as 1 milkborne human infection per 6,410,256 servings (with mean of 1 infection per 334,455 servings). Applying this risk to an assumed 583 million annual raw milk servings consumed results in a median of 91 infections per year (IQR: 12-636; mean: 1,743). Note that no literature identified could parameterize the proportion of raw milk servings arising from farmstore versus retail purchase, and thus, this projection was made under the assumption that all raw milk servings consumed are from the farmstore pathway.

### 3.3. Sensitivity Analysis

Tornado plots are given in **Figures 4a-c**. In both models, the viral titer in infected cow milk *V_M_* was very strongly correlated with the p(infection) (pasteurized: =0.94; raw, farmstore: =0.89; raw, retail: =0.85). This metric was also sensitive to the US herd-level H5N1 prevalence *p_nat’l_* in both models (pasteurized: =0.80; raw, farmstore: =0.69; raw, retail: =0.60).

In the pasteurized milk model, parameter *p_herd_* (within-herd H5N1 prevalence) demonstrated moderate positive correlation ( =0.52). Other parameters (*Y_I_* and *H_Z_*) yielded weak or negligible correlation. In the raw milk model, weak positive correlation was observed in *p_herd_* (farmstore: =0.33; retail: =0.27). For the farmstore purchase pathway, moderate negative correlations were observed for the duration *U_F_* ( =-0.63) and temperature *T_F_*( =-0.56) of farmstore storage, and the duration *U_SC_* ( =-0.50) and temperature *T_SC_* ( =-0.45) of storage at the consumer residence. Similarly, in the retail purchase pathway, the duration *U_SR_*( =-0.55) and temperature *T_SR_* ( =-0.43) of storage at retail display moderate negative correlation. Infected cow relative milk yield *Y_I_* was negligible in both purchasing pathways and herd size *H_W_* was weak in both (farmstore: =-0.14; retail: =-0.11). Compared against their counterparts in the pasteurized milk model, the strengths and directions of statistically significant parameters were comparable with the exceptions of temperature and storage parameters, which failed to achieve significance in the pasteurized milk model. This is likely due to the increased relative importance of product spoilage and discard in reducing risk in the raw milk model compared with the pasteurized milk model.

### 3.4. Scenario Analysis

Increases in the pasteurization efficacy parameter *L* produced a progressively lower median p(infection) at each level tested. The median p(infection) values were 5.01E-14 and 4.70E-20, and mean p(infection) values were 1.65E-13 and 1.26E-19 in the 6-log and 14-log reduction pasteurization scenarios, respectively (**Figure 5** and **Table S2**).

**FIGURE 5.**
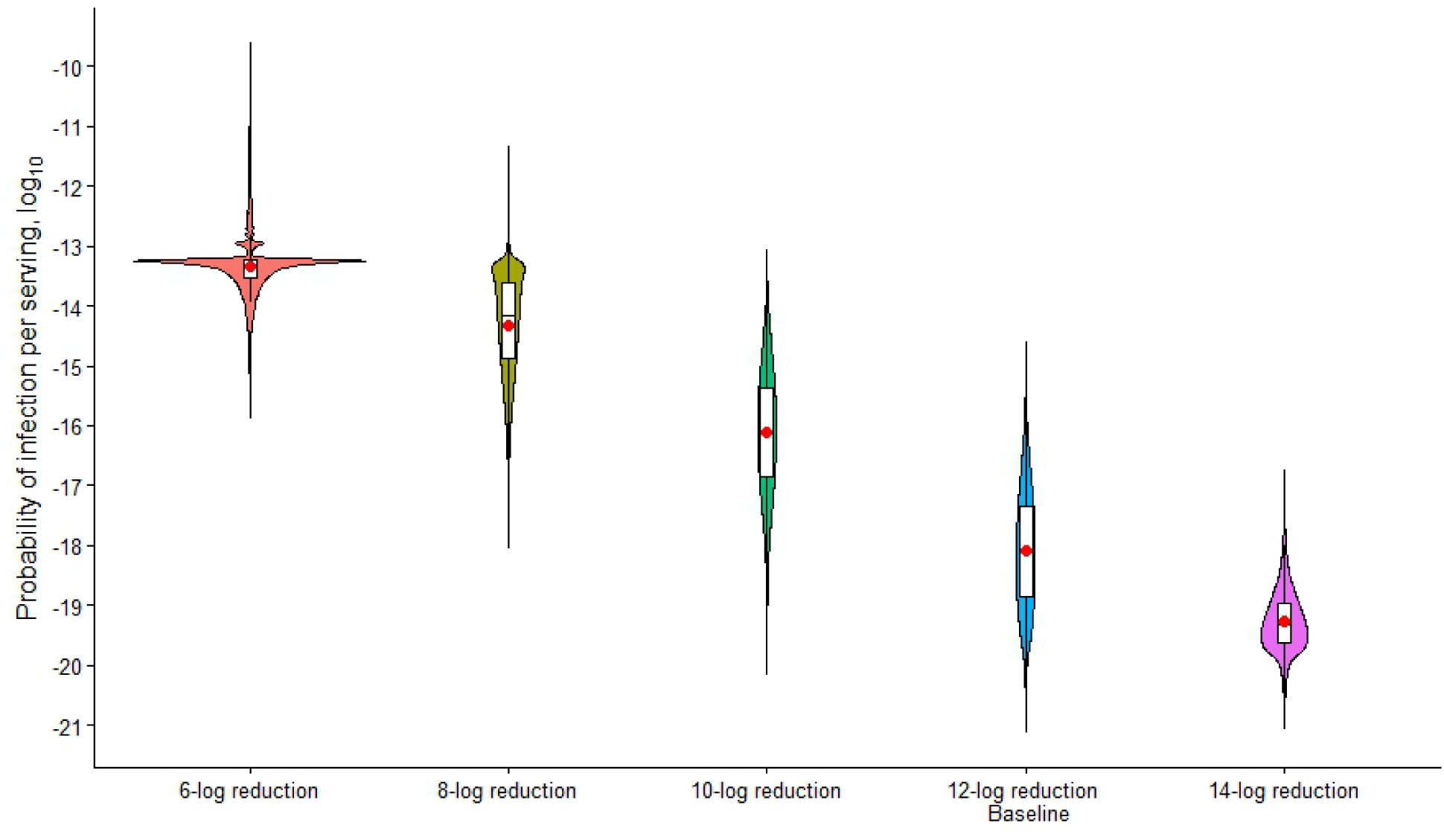
Violin plots for the scenario analysis of the impact of varying pasteurization log reduction parameter *L* on the probability of H5N1 infection per pasteurized milk serving (p(infection)). For the purposes of visualization, a logarithmic (base-10) Y-axis is employed. The mean is denoted with the red dot.

In the raw milk supply chain, bulk tank PCR testing was more effective than improved animal diversion in numerically reducing p(infection). For all three levels of diversion, the addition of bulk tank PCR testing reduced both the mean and median p(infection) 100-fold (**Figures 6** and **S1**). In the farmstore purchase pathway (**Table S3**), without bulk tank PCR, increasing infected cow diversion over a 25% baseline to 50% reduced the median (mean) p(infection) from 1.56E-7 (2.99E-6) to 1.06E-7 (2.08E-6); with the addition of bulk tank PCR, these values at 25% diversion was 2.49E-9 (4.78E-8), then 1.69E-9 (3.33E-8) at 50% diversion. Results for the retail purchase pathway were similar (**Table S4**). An analysis of a hypothetical improvement in the diagnostic (i.e. increase in *Se* from 98.4% to 99.9%) and/or analytical (i.e. decrease in *LoD* from 1.5 to 1.0 logTCID_50_/0.1mL) sensitivity of this PCR test demonstrated that the higher *Se* results in a 1-log decrease in median (mean) farmstore p(infection) to 1.55E-10 (2.99E-9), while the lower limit of detection confers no appreciable numerical reduction in risk (**Analysis S1**, **Table S6**, and **Figure S2**). **Analysis S4** demonstrated that in the absence of bulk tank PCR testing, even at 99.9% diversion, the median p(infection) of farmstore-purchased milk is still numerically higher than that of baseline diversion with PCR surveillance (**Table S9**).

**FIGURE 6.**
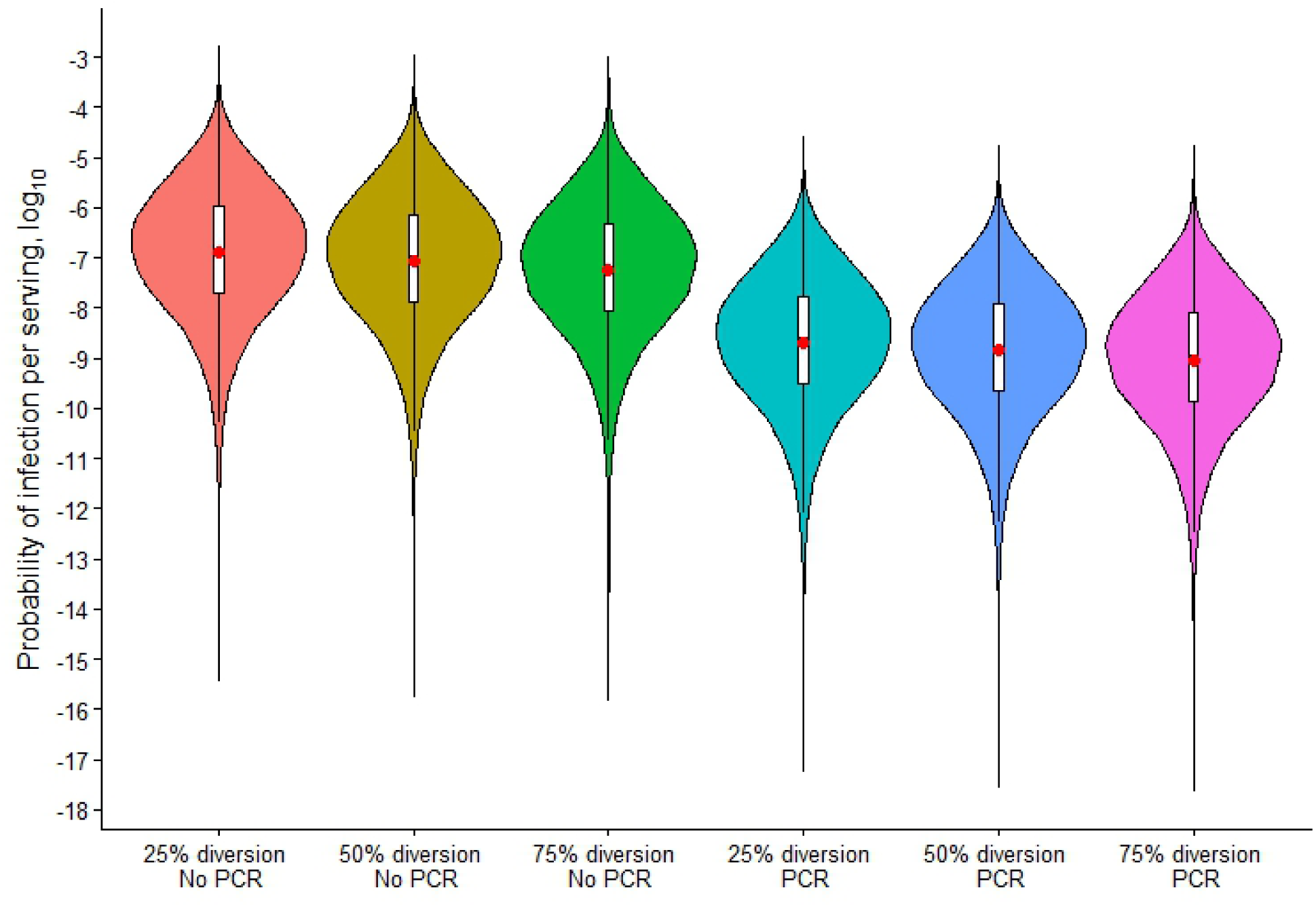
Violin plots for the scenario analysis of bulk tank milk PCR testing (defined by parameters *Se*, *Sp* and *LoD*) and improved infected cow diversion (parameter *D*) on the probability of infection per farmstore-purchased raw milk serving (p(infection)). For the purposes of visualization, a logarithmic (base-10) Y-axis is employed. The mean is denoted with the red dot.

### 3.5. Uncertainty Analysis

Accounting for the uncertainty in the dose-response parameter *r* according to the ranges reported by Chen et al. (2025) results in largely negligible (0 to 1-log shifts) increases in p(infection) and a slightly wider distribution of the risk compared with the baseline models (**Table S5** and **Figure 7**). The effect of uncertainty in this parameter was consistent in both raw and pasteurized milk, and across both raw milk purchasing pathways.

**FIGURE 7.**
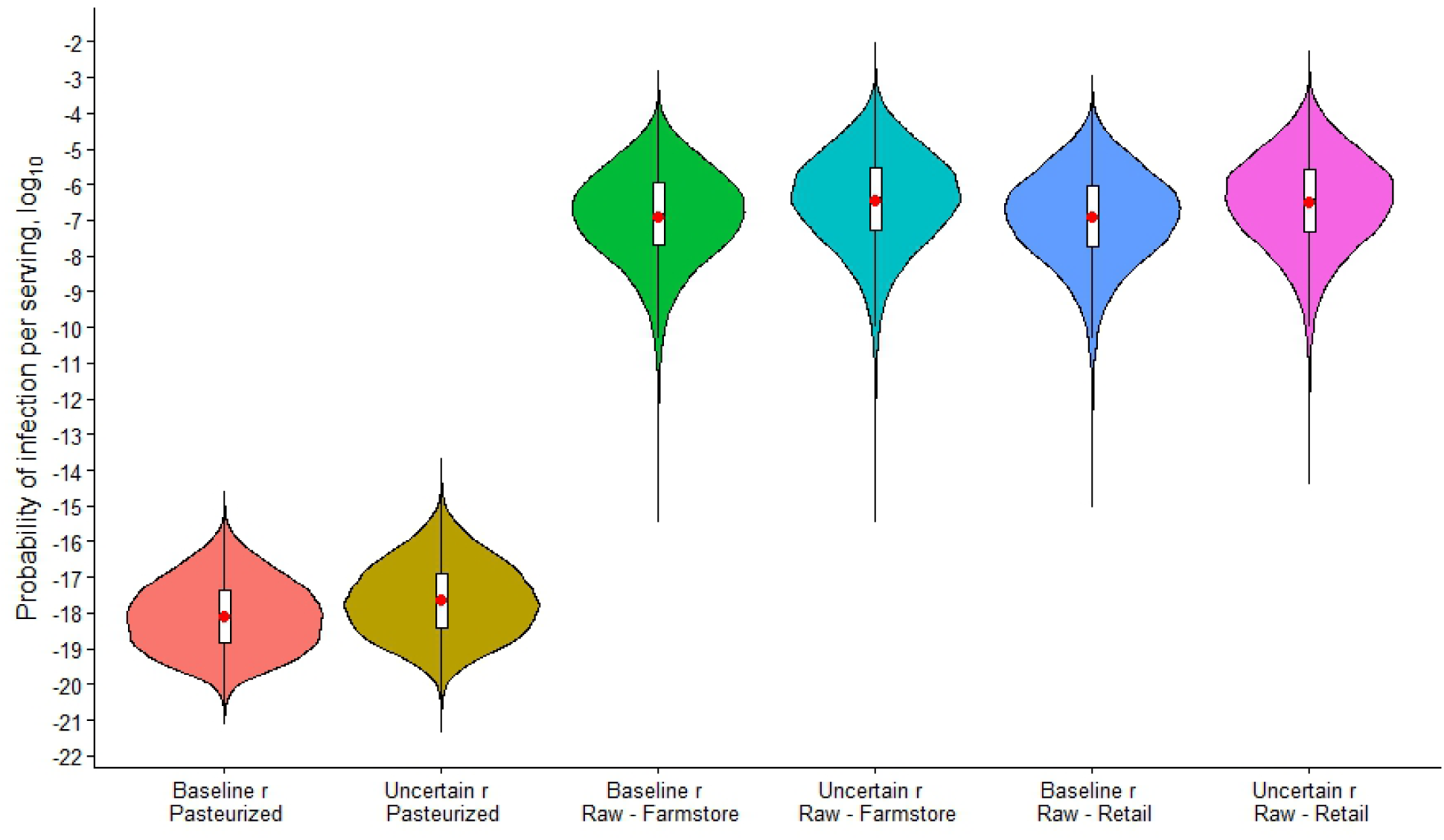
Violin plots for the analysis of the impact of uncertainty in dose-response parameter *r* on the probability of infection per serving of pasteurized or farmstore- or retail-purchased raw milk. For the purposes of visualization, a logarithmic (base-10) Y-axis is employed. The mean is denoted with the red dot.

### 3.6. Assumption Analysis

As seen in **Figure 8** and **Table 2**, use of empirical herd infection counts under the assumption of geographic homogeneity rather than clustering in the distribution of affected herds results in an approximately 1-log decrease in p(infection) for pasteurized milk. This results from a truncation in the distribution in the *K* parameter, which took a maximum value of 192 in the empirical data and 4,924 when calculated from state-level prevalences. While the numerical impact of this assumption on the broader conclusions drawn from this study is limited, it is worth noting that the constituent mechanistic assumption that all infected US herds are equally likely to ship milk to the modeled processing plant is not realistic due to interstate milk shipping regulations.

**FIGURE 8.**
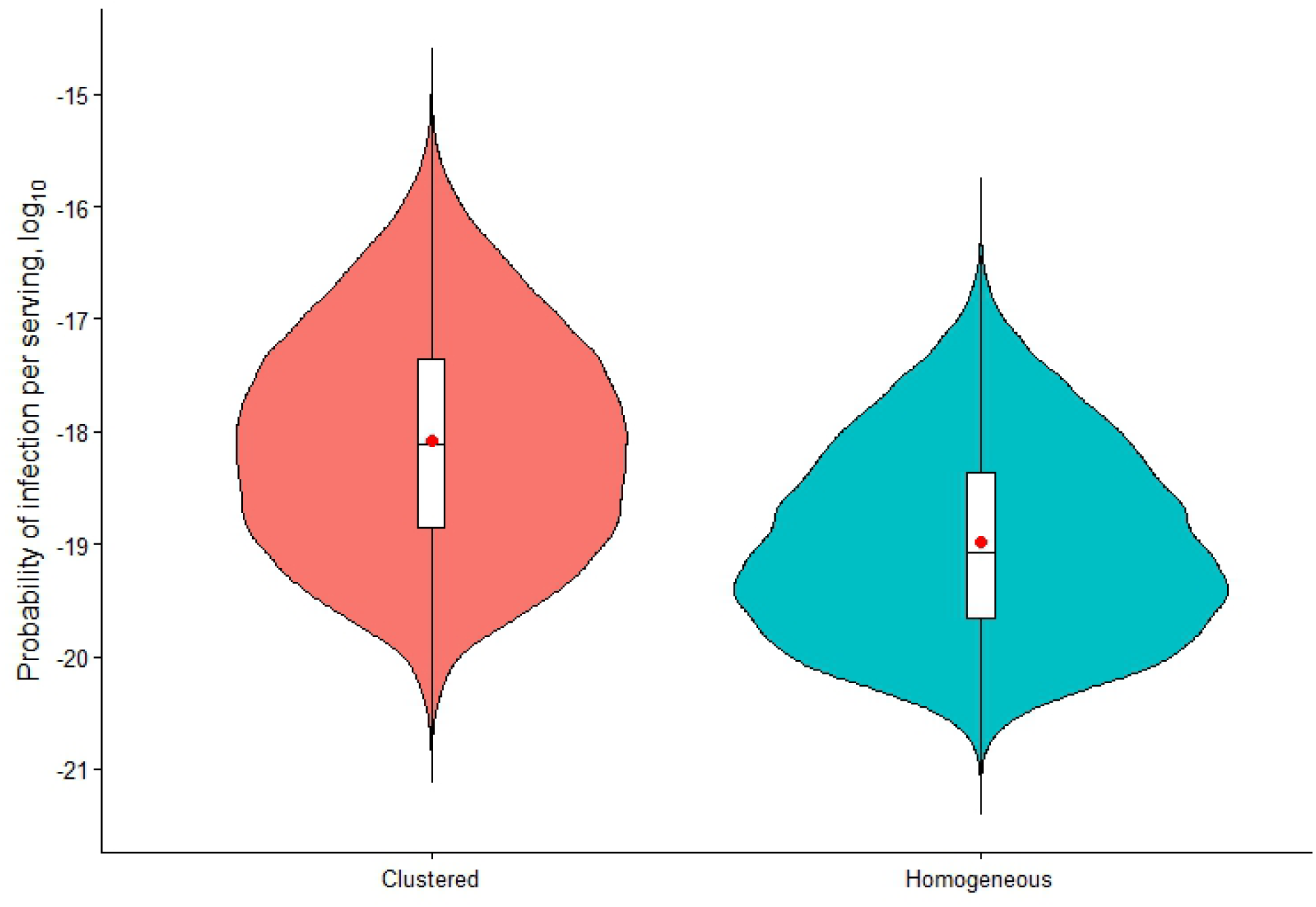
Violin plots for the probability of H5N1 infection per pasteurized milk serving (p(infection)) calculated with two different methods for parameterizing the number of infected herds in the US (*K*). The clustered approach indirectly accounts for the clustering phenomenon of localized outbreaks amongst herds in a geographic area. The homogeneous approach assumes a uniform geographic distribution of infected herds in the US. For the purposes of visualization, a logarithmic (base-10) Y-axis is employed. The mean is denoted with the red dot.

**TABLE 2.** is in landscape format in *Supplementary Materials*.

Summary statistics for p(infection) resulting from analyses of **Assumptions 3** and **4** are given in **Tables 3** and **4**, respectively. These respectively demonstrate that the impact of assuming uniform viral distribution in fluid suspension and equitable *p_inf_* between raw and conventional herds on the conclusions of our risk analysis is minimal.

**TABLE 3.**
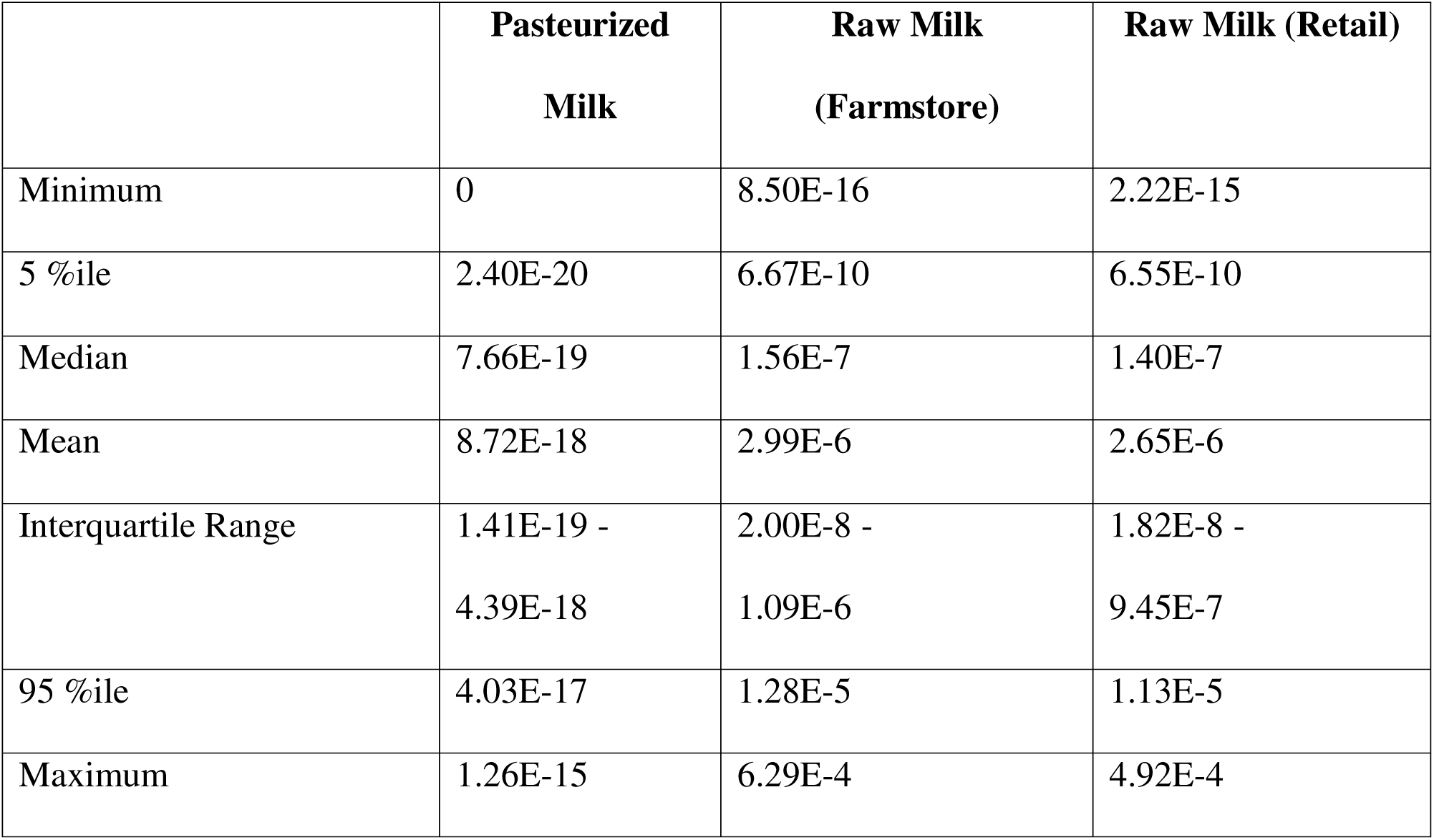
Summary statistics for the probability of H5N1 infection per serving (p(infection); unitless) of pasteurized or raw fluid milk when viral particles are assumed to cluster in suspension.

|  | **Pasteurized**<br>**Milk** | **Raw Milk**<br>**(Farmstore)** | **Raw Milk (Retail)** |
| --- | --- | --- | --- |
| Minimum | 0 | 8.50E-16 | 2.22E-15 |
| 5 %ile | 2.40E-20 | 6.67E-10 | 6.55E-10 |
| Median | 7.66E-19 | 1.56E-7 | 1.40E-7 |
| Mean | 8.72E-18 | 2.99E-6 | 2.65E-6 |
| Interquartile Range | 1.41E-19 -<br>4.39E-18 | 2.00E-8 -<br>1.09E-6 | 1.82E-8 -<br>9.45E-7 |
| 95 %ile | 4.03E-17 | 1.28E-5 | 1.13E-5 |
| Maximum | 1.26E-15 | 6.29E-4 | 4.92E-4 |

**TABLE 4.** Summary statistics for the probability of H5N1 infection per serving (p(infection); unitless) of raw fluid milk when raw milk herds are 50% less or more likely to experience H5N1 infection.

|  | **Baseline** |  | **50% Less Likely** |  | **50% More Likely** |  |
| --- | --- | --- | --- | --- | --- | --- |
|  | **Farmstore** | **Retail** | **Farmstore** | **Retail** | **Farmstore** | **Retail** |
| Minimum | 8.50E-16 | 2.22E-15 | 4.25E-16 | 1.11E-15 | 1.27E-15 | 3.33E-15 |
| 5 %ile | 6.67E-10 | 6.65E-10 | 3.34E-10 | 3.32E-10 | 1.00E-9 | 9.97E-10 |
| Median | 1.56E-7 | 1.40E-7 | 7.78E-8 | 7.00E-8 | 2.33E-7 | 2.10E-7 |
| Mean | 2.99E-6 | 2.65E-6 | 1.49E-6 | 1.33E-6 | 4.48E-6 | 3.98E-6 |
| Interquartile Range | 2.00E-8 - 1.09E-6 | 1.82E-8 - 9.45E-7 | 1.00E-8 - 5.44E-7 | 9.08E-9 - 4.72E-7 | 3.00E-8 - 1.63E-6 | 2.72E-8 - 1.42E-6 |
| 95 %ile | 1.28E-5 | 1.13E-05 | 6.40E-6 | 5.63E-6 | 1.92E-5 | 1.69E-5 |
| Maximum | 6.29E-4 | 4.52E-4 | 3.15E-4 | 2.23E-4 | 9.44E-4 | 6.78E-4 |

## 4. DISCUSSION

Irrespective of the mode of human exposure, whether foodborne or otherwise, H5N1 viruses remain a public health concern because of their intrinsically high propensity for genetic mutation or reassortment in an infected host, which heightens the risk that the virus could acquire the ability for sustained human to human transmission (Lin et al., 2024). This QMRA predicts the risk of human H5N1 infection from consumption of pasteurized fluid milk to be extremely low (**Figure 3** and **Table S1**). According to research surrounding H5N1 and HTST pasteurization, 12-log reduction is highly efficacious at reducing the public health risk (Spackman et al., 2024a). Still, demonstration of pasteurization efficacy is fundamentally limited by the viral concentration of the initial sample (Nooruzzaman et al., 2025a). To definitively demonstrate the efficacy of pasteurization, assays with supraphysiological concentrations of virus must be conducted. Pasteurization remains the most effective method for reducing risk amidst this H5N1 outbreak. Scenario analysis (**Table S2**) demonstrates how, should pasteurization efficacy be overestimated or a pasteurization failure go undetected, the risk to public health would be increased A consequence of the exponential dose-response model utilized in this study is the inherent assumption that a single infectious organism is probabilistically capable of establishing infection in the host (Nilsen and Wyller, 2016). Quantification of the low-range of the exponential dose-response model is inherently uncertain since, at very low doses, the probability of infection is naturally small, and a prohibitively large sample size would be required to quantify it with the necessary level of confidence (Nilsen and Wyller, 2016).

However, with contaminated pasteurized servings containing only 1-2 TCID_50_, and the dose-response parameter *r* of Chen et al. (2025) adjusted to account for a 3-log microbial clearance through gastric passage, the probability of infection is functionally negligible: even with 100-fold uncertainty in parameter *r* in either direction. However, it must be noted that, while the authors derived this equation from an established human influenza A inhalational dose-response curve and exposure route extrapolation was informed by published literature, the lack of a validated orogastric dose-response curve for human H5N1 infection introduces significant uncertainty into the risk projections.

It is also important to note that, to date, none of the reported human H5N1 cases have been positively attributed to consumption of contaminated dairy products. The absence of a reported foodborne H5N1 case may be a result of (i) subclinical infection, (ii) non-reporting of clinical infections, (iii) effectiveness of the implemented control strategies, and/or (iv) inability of H5N1 clade 2.3.4.4b to infect humans via ingestion.

As previously discussed, a significant body of literature substantiates the possibility of H5N1 infection by ingestion, demonstrating it definitively in animal models and providing evidence to support the plausibility of this phenomenon in humans. In their 2013 review, Killingley and Nguyen-van-Tam express the role of animal models in the evidence base of influenza research, while still maintaining that the findings of such studies in relation to human pathophysiology is “uncertain and scientifically challengeable.” In the Rosenke et al. (2025) experiment, macaques, noted to be “widely used surrogate models for human infectious diseases,” did not display clinical signs despite undergoing seroconversion and showing histopathological evidence of localized infection. This may be interpreted in the context of wastewater surveillance of H5N1 clade 2.3.4.4b (Tisza et al., 2024). Detection of H5N1 in Texas wastewater coincided with the emergence of the disease syndrome in dairy cattle and predates the official announcement of the causative agent. While genetic analysis of wastewater samples indicates primarily bovine contribution (likely milk effluent from farms or processing plants), viral shedding in human sewage cannot be ruled out (Honein et al., 2024).

Furthermore, it is unknown what disease syndrome a foodborne H5N1 infection would produce in a symptomatic human. Case reports and epidemiological data from the outbreak thus far suggest a predominantly mild conjunctival or upper respiratory illness, likely not severe or concerning enough to merit seeking treatment. In laboratory animal studies, clinical signs after H5N1 ingestion ranged from none to weight loss and lethargy, up to and including mortality (Halwe et al., 2024; Rosenke et al., 2025; Lipatov et al., 2009; Guan et al., 2024; Eisfeld et al., 2024; Gu et al., 2024). It is thought that viral contact with the oropharynx during deglutition is the route of entry sufficient to establish infection in these models. While such a route of transmission is previously undescribed, as in the review by Killingley and Nguyen-van-Tam (2013), it is mechanistically possible given its similarity to droplet transmission. Perhaps presentation with conjunctivitis in humans is a function of the route of exposure, including the splash of contaminated material into the mucous membranes of the face. Other routes of exposure may produce a different array of symptoms. Additionally, Garg et al. (2025) report that only 38% of the H5N1 clade 2.3.4.4b human cases in their study sought medical care. Therefore, the absence of a documented case attributable to milk consumption may be a result of asymptomaticity or under-detection. Using the baseline model in Morris et al. (2024), which assumes that the severity of H5N1 infections is similar to seasonal influenza and uses parameters for urgent care settings, the median probability of detecting an influenza infection through this system is 1.29% (5-95%ile: 0.66% - 2.36%). Employing the top-down approach by Chen et al. (2025), and after adjusting for the differences in the serving size between our and their QMRA, the mean minimal incidence risk (i.e., average p(infection) that would result in a 95% probability of observing at least one H5N1 case attributable to milk consumption) is 3.44E-08 (5-50-95%ile: 3.82E-09 - 1.05E-08 - 1.03E-07) for pasteurized milk and 2.83E-06 (5-50-95%ile: 3.15E-07 – 8.61E-07 – 8.50E-06) for raw milk. By contrast, the mean p(infection) from pasteurized milk from our model is 8.71E-18 (5-50-95%ile: 2.39E-20 - 7.66E-19 - 4.02E-17). For farmstore- purchased raw milk, the predicted mean p(infection) is 2.99E-6 (5-50-95%ile: 6.67E-10 - 1.56E- 7 - 1.28E-5). Next, by multiplying the predicted mean p(infection) with the number of respective servings (pasteurized or raw) consumed per annum and the probability of detecting an influenza infection (mentioned above; Morris et al, 2024), we can predict the number of confirmed cases attributed to yearly milk consumption. Thus, in a year in the USA, we would expect an average of 7.18E-09 (5-95%ile: 1.35E-09 - 1.78E-08) confirmed cases for pasteurized milk, supporting that a confirmed outbreak from pasteurized milk is highly unlikely. Using the same approach, we would expect an average of 29.93 (5-95%ile: 5.61 - 74.18) confirmed cases for raw milk. Given the small expected number of detected illness cases predicted for the raw exposure pathway, under the fundamental assumption of the model that H5N1 is infectious to humans via ingestion, it is plausible that an outbreak from raw milk would go unrecognized. Note that the timeframe for this analysis is restricted to one year due to the use of the early-outbreak USDA data, after which the implementation of public health mandates confounded the natural course of the outbreak and risk projections would no longer be representative.

A third potential contributing factor to the absence of reported foodborne H5N1 infections in people could be the efficacy of the aforementioned Federal Order control strategies, including mandatory animal and milk testing (USDA, 2024). Results of this milk monitoring have demonstrated markedly decreased recovery rates of IAV (Tarbuck et al., 2025). As of 8 February 2026, the USDA-APHIS dashboard reports that no new detections of H5N1 have been reported in cattle in the past 30 days.

Evidence against the possibility of H5N1 infection by ingestion must also be considered. The affinity of bovine H5N1 viruses for human-type α2,6 sialic acid receptors relative to avian α2,3 conformations was questioned early in the outbreak (Eisfeld et al., 2024). *In vitro* studies examining preferential binding of bovine isolates to these two varieties of receptors have yielded conflicting results (Chopra et al., 2025; Santos et al., 2025). The dubious affinity for bovine isolates to bind human-type receptors is implicated in the ability (or inability) of H5N1 clade 2.3.4.4b to infect people via ingestion. For consideration, in our raw milk model, under the assumptions of human foodborne infectivity, at least 1 foodborne infection was projected in >99.9% of iterations for the farmstore and retail pathways: values that make the absence of detection probabilistically unlikely if infection is possible and symptomatic. As the outbreak continues towards resolution, the absence of definitive foodborne human cases should be acknowledged with due consideration given to the quality and quantity of scientific evidence supporting the presence of this hazard.

We also encourage careful scrutiny of the limitations and assumptions of our models (*Supplementary Materials*), and acknowledge the inherent interspecies and inter-route extrapolation of our supporting evidence and dose-response model, respectively. Cumulatively, the assumptions of clustered distributions of infected herds, uniform spatial dispersion of viruses in fluid suspension, and equal probability of herd infection for conventional versus raw milk operations introduce negligible-to-low levels of uncertainty into our model. Also, our model does not account for cumulative risk from consumption of multiple subinfectious doses over time nor does it differentiate between milkfat levels. Repeated exposure over time may alter susceptibility or result in sensitization, violating the assumption of inter-dose independence. No empirical data from repeated human exposure are available to support dose response modeling; however, in Nooruzzaman et al. (2025b), repeated ingestion of contaminated milk led to H5N1 infection in ferrets, supporting the need for further research in this phenomenon. Furthermore, if virions distribute differently in fat globules versus the liquid fraction, the true risk may be shifted in either direction. As Spackman et al. report slight differences in the concentration of viral material amongst different milkfat levels (2024b), continued surveillance is required to determine if this is a function of processing or sample size.

Nevertheless, the public health burden from pasteurized milk predicted by this QMRA is extremely low, in that no iterations produced projected foodborne infections. Note that this output is calculated with per annum consumption data and thus represents the projected *annual* number of infections utilizing epidemiologic data collected between March and November of 2024; seeing as initiatives to mitigate the outbreak have resulted in a reduced monthly incidence of herd infection over time, extrapolating data from the early phases of the outbreak to an entire year means our projections likely capture the upper limit of true risk.

Ingestion of raw milk carries an inherent danger of foodborne illness (Lando et al., 2022; CDC, 2024b; FDA, 2024b, Koski et al., 2022). Our model demonstrates that, without pasteurization, the p(infection) is dramatically higher. Given the popularity of raw milk amongst US consumers, in light of this outbreak there is an urgent need for new technological, educational, and policy solutions to protect the health of raw milk consumers. Here we have demonstrated the efficacy of two interventions. PCR testing of raw milk herd bulk tanks is efficacious in numerically reducing p(infection), more so than our representation of improved identification and diversion of milk from infected cows. However, despite its comparatively weaker effect, diverting infected cows is still important, as this relates to sensitivity in both models to viral titers in infected cow milk (*V_M_*). The practical ability to achieve an appreciable or significant reduction in p(infection) from diversion of infected cows alone will be determined by labor, diagnostic, economic, and logistic constraints on any given operation; we show in **Analysis S4** that the necessary level of detection cannot be feasibly attained on most farms, exceeding the upper limit of 75% set by the authors as a realistic maximum. It is also possible that the development of improved molecular diagnostics will lead to better sensitivity, specificity, and limits of detection that will increase the risk mitigation value of these technologies in bulk tank milk testing. The potential impact of such a diagnostic test is briefly examined in **Analysis S1**. The USDA is currently implementing a bulk tank milk testing surveillance plan at the state and locoregional levels (USDA-APHIS, 2024a). This will likely be more important for smaller herds, as the brief **Analysis S3** demonstrates that p(infection) is numerically, but not practically, lower when the maximum of herd size parameter *H_W_* is raised to account for larger commercial raw milk operations. Also in sensitivity analysis, the decreased H5N1 infection risk in raw milk associated with increased storage temperatures/times is a function of viral decay and milk spoilage; excessive or inappropriate storage of dairy products is not advisable due to the risk of illness from other foodborne pathogens that may be present in raw milk.

Additional information is needed to make a QMRA accounting for all dairy products. As research characterizing H5N1 in other dairy foods becomes available, the framework of our model may be adapted. For example, academics and industry stakeholders have expressed interest in studying H5N1 in raw milk cheese. The expansion of our models to fill this knowledge gap is an area of active investigation for the authors.

In conclusion, based on the results of this QMRA and in accordance with the ACMSF qualitative criteria (2012), we find the risk of human H5N1 infection from consumption of pasteurized fluid cow’s milk to be **negligible**: characterized by **moderate uncertainty**. Due to the absence of pasteurization, the risk for raw milk is demonstrably higher but remains **low** with **moderate uncertainty**. Additionally, the absence of reported clinical cases indicates **low severity** for both supply chains. Consumption of raw milk has historically been discouraged due to the risk of foodborne illness; in the context of this outbreak, the emergence of a novel potential milkborne pathogen emphasizes both the risk of this practice and the need for solutions to effectively protect the health of raw milk consumers. This risk may be reduced with implementation of risk mitigation interventions, including thermal treatment and bulk tank PCR testing. As new information becomes available, the models are subject to refinement.

## Data Availability

All data produced in this work are available upon reasonable request to the corresponding author.

https://github.com/IvanekLab/H5N1-Dairy-QRA

## Data Availability

All data produced in this work are available upon reasonable request to the corresponding author.

https://github.com/IvanekLab/H5N1-Dairy-QRA

## ACKNOWLEDGEMENTS

Research reported in this publication was supported by the Office of the Director, National Institutes of Health of the National Institutions of Health (NIH) under Award Number T32ODO011000 to KK. The content is solely the responsibility of the authors and does not necessarily represent the official views of the National Center for Research Resources or the NIH. Additionally, this research was partially supported by grants to RI from the National Institute of Food and Agriculture, USDA, Hatch under Accession Number 7000433, as well as Multistate Research Funds Accession Number 1016738.

## DATA AVAILABILITY STATEMENT

The models are available at https://github.com/IvanekLab/H5N1-Dairy-QRA.

