## Supplementary materials for "Quantitative microbial risk assessment of human H5N1 infection from consumption of fluid cow’s milk"

**TABLE S1 | Summary statistics for key model outputs of a quantitative microbial risk assessment (QMRA) of human H5N1 infection from consumption of raw or pasteurized fluid cow’s milk**. Two purchasing pathways (via farmstore and via retail) for raw milk are differentiated. *Note: 0.00 logTCID_50_ = 1 TCID_50_ (“log” indicates log_10_ transformation)

|  | **Pasteurized Milk Model** | | | **Raw Milk Model** | | | | | |
| --- | --- | --- | --- | --- | --- | --- | --- | --- | --- |
|  |  | | | **Farmstore Purchasing Pathway** | | | **Retail Purchasing Pathway** | | |
|  | Probability of Infection per Serving  (p(infection)) (unitless) | Quantity of Virus per Contaminated Serving  (Q_serv_) (logTCID_50_) | Probability of Serving Contamination  (p(serving))  (unitless) | Probability of Infection per Serving  (p(infection))  (unitless) | Quantity of Virus per Contaminated Serving  (Q_serv_) (logTCID_50_) | Probability of Serving Contamination  (p(serving))  (unitless) | Probability of Infection per Serving  (p(infection))  (unitless) | Quantity of Virus per Contaminated Serving  (Q_serv_) (logTCID_50_) | Probability of Serving Contamination  (p(serving))  (unitless) |
| Minimum | 1.51E-21 | 0.00* | 1.58E-9 | 8.50E-16 | 3.04 | 8.03E-4 | 2.22E-15 | 3.09 | 8.03E-4 |
| 5 %ile | 2.39E-20 | 0.00 | 2.43E-8 | 6.67E-10 | 5.19 | 3.21E-3 | 6.65E-10 | 5.16 | 3.21E-3 |
| Median | 7.66E-19 | 0.00 | 7.67E-7 | 1.56E-7 | 6.86 | 3.13E-2 | 1.40E-7 | 6.81 | 3.13E-2 |
| Mean | 8.71E-18 | 0.019 | 8.66E-6 | 2.99E-6 | 6.85 | 4.00E-2 | 2.65E-6 | 6.81 | 4.00E-2 |
| Interquartile Range | 1.41E-19 - 4.39E-18 | 0.00 - 0.00 | 1.41E-7 - 4.41E-6 | 2.00E-8 - 1.09E-6 | 6.13 - 7.58 | 1.40E-2 - 5.78E-2 | 1.82E-8 - 9.45E-7 | 6.09 - 7.53 | 1.40E-2 - 5.78E-2 |
| 95 %ile | 4.02E-17 | 0.30 | 4.03E-5 | 1.28E-5 | 8.47 | 1.07E-1 | 1.13E-05 | 8.42 | 1.07E-1 |
| Maximum | 1.25E-15 | 0.30 | 1.32E-3 | 6.29E-4 | 9.69 | 2.14E-1 | 4.52E-4 | 9.81 | 2.14E-1 |

**TABLE S2 | Summary statistics for the scenario analysis of the impact of varying pasteurization log (base-10) reduction parameter *L* on the probability of H5N1 infection per pasteurized milk serving (p(infection)).**

|  | **6-log** | **8-log** | **10-log** | **12-log** (Baseline) | **14-log** |
| --- | --- | --- | --- | --- | --- |
| Minimum | 1.54E-16 | 1.57E-18 | 1.42E-20 | 1.51E-21 | 1.19E-21 |
| 5 %ile | 7.57E-15 | 1.30E-16 | 1.28E-18 | 2.39E-20 | 1.04E-20 |
| Median | 5.01E-14 | 6.68E-15 | 7.89E-17 | 7.66E-19 | 4.70E-20 |
| Mean | 1.65E-13 | 1.53E-14 | 7.53E-16 | 8.71E-18 | 1.26E-19 |
| Interquartile Range | 3.03E-14 - 5.83E-14 | 1.33E-15 - 2.40E-14 | 1.37E-17 - 4.40E-16 | 1.41E-19 - 4.39E-18 | 2.41E-20 - 1.08E-19 |
| 95 %ile | 3.55E-13 | 5.21E-14 | 3.78E-15 | 4.02E-17 | 4.47E-19 |
| Maximum | 2.07E-10 | 2.57E-12 | 4.12E-14 | 1.25E-15 | 1.28E-17 |

**TABLE S3 | Summary statistics for the probability of H5N1 infection per raw milk serving (p(infection)) obtained through farmstore purchase in the scenario analysis of the impact of bulk tank milk PCR testing (defined by parameters *Se*, *Sp* and *LoD*) and improved infected cow diversion (parameter *D*).**

|  | **No PCR Testing** | | | **PCR Testing** | | |
| --- | --- | --- | --- | --- | --- | --- |
|  | **25% Diversion** (Baseline) | **50% Diversion** | **75% Diversion** | **25% Diversion** | **50% Diversion** | **75% Diversion** |
| Minimum | 8.50E-16 | 4.35E-16 | 3.72E-16 | 1.35E-17 | 6.87E-18 | 5.67E-18 |
| 5 %ile | 6.67E-10 | 4.49E-10 | 2.91E-10 | 1.06E-11 | 7.08E-12 | 4.57E-12 |
| Median | 1.56E-7 | 1.06E-7 | 6.98E-8 | 2.49E-9 | 1.69E-9 | 1.11E-9 |
| Mean | 2.99E-6 | 2.08E-6 | 1.40E-6 | 4.78E-8 | 3.33E-8 | 2.23E-8 |
| Interquartile Range | 2.00E-8 - 1.09E-6 | 1.35E-8 - 7.39E-7 | 8.91E-9 - 4.90E-7 | 3.19E-10 - 1.74E-8 | 2.14E-10 - 1.18E-8 | 1.41E-10 - 7.84E-9 |
| 95 %ile | 1.28E-5 | 8.83E-6 | 5.89E-6 | 2.05E-7 | 1.41E-7 | 9.43E-8 |
| Maximum | 6.29E-4 | 4.34E-4 | 4.14E-4 | 1.01E-5 | 6.94E-6 | 6.63E-6 |

**TABLE S4 | Summary statistics for the probability of H5N1 infection per raw milk serving (p(infection)) obtained through retail purchase in the scenario analysis of the impact of bulk tank milk PCR testing (defined by parameters *Se*, *Sp* and *LoD*) and improved infected cow diversion (parameter *D*).**

|  | **No PCR Testing** | | | **PCR Testing** | | |
| --- | --- | --- | --- | --- | --- | --- |
|  | **25% Diversion** (Baseline) | **50% Diversion** | **75% Diversion** | **25% Diversion** | **50% Diversion** | **75% Diversion** |
| Minimum | 2.22E-15 | 1.53E-15 | 7.93E-16 | 3.49E-17 | 2.38E-17 | 1.21E-17 |
| 5 %ile | 6.65E-10 | 4.43E-10 | 2.94E-10 | 1.05E-11 | 6.98E-12 | 4.61E-12 |
| Median | 1.40E-7 | 9.49E-8 | 6.24E-8 | 2.24E-9 | 1.52E-9 | 9.96E-10 |
| Mean | 2.65E-6 | 1.84E-6 | 1.23E-6 | 4.25E-8 | 2.95E-8 | 1.98E-8 |
| Interquartile Range | 1.82E-8 - 9.45E-7 | 1.22E-8 - 6.42E-7 | 8.06E-9 - 4.31E-7 | 2.90E-10 - 1.51E-8 | 1.95E-10 - 1.03E-8 | 1.28E-10 - 6.89E-9 |
| 95 %ile | 1.13E-05 | 7.76E-6 | 5.16E-6 | 1.80E-7 | 1.24E-7 | 8.25E-8 |
| Maximum | 4.52E-4 | 3.16E-4 | 2.17E-4 | 7.23E-6 | 5.06E-6 | 3.48E-6 |

**FIGURE S1 |** **Violin plots for the scenario analysis of the impact of bulk tank milk PCR testing (defined by** **parameters *Se*, *Sp* and *LoD*) and improved infected cow diversion (parameter *D*) on probability of infection per raw milk serving (p(infection)) obtained through retail purchase.** For the purposes of visualization, a logarithmic (base-10) Y-axis is employed. The mean is denoted with the red dot.

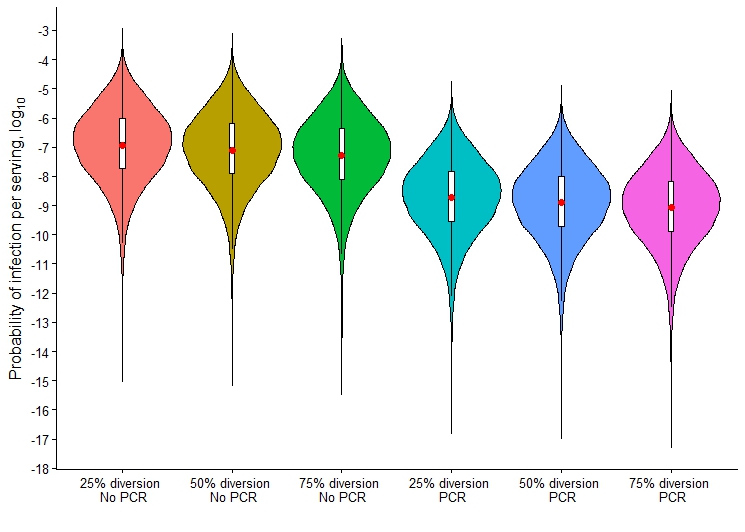

**TABLE S5 | Summary statistics for the analysis of the impact of uncertainty in dose-response parameter *r* on the probability of infection per pasteurized or farmstore- or retail-purchased raw milk.**

|  | **Pasteurized Milk Model** | | **Raw Milk Model** | | | |
| --- | --- | --- | --- | --- | --- | --- |
|  |  | | **Farmstore Purchasing Pathway** | | **Retail Purchasing Pathway** | |
|  | **Baseline *r*** | **Uncertain *r*** | **Baseline *r*** | **Uncertain *r*** | **Baseline *r*** | **Uncertain *r*** |
| Minimum | 1.51E-21 | 9.24E-22 | 8.50E-16 | 8.38E-16 | 2.22E-15 | 9.64E-15 |
| 5 %ile | 2.39E-20 | 5.11E-20 | 6.67E-10 | 1.61E-9 | 6.65E-10 | 1.60E-9 |
| Median | 7.66E-19 | 2.06E-18 | 1.56E-7 | 4.24E-7 | 1.40E-7 | 3.81E-7 |
| Mean | 8.71E-18 | 3.07E-17 | 2.99E-6 | 9.73E-6 | 2.65E-6 | 8.50E-6 |
| Interquartile Range | 1.41E-19 - 4.39E-18 | 3.66E-19 - 1.28E-17 | 2.00E-8 - 1.09E-6 | 5.05E-8 - 3.11E-6 | 1.82E-8 - 9.45E-7 | 4.60E-8 - 2.71E-6 |
| 95 %ile | 4.02E-17 | 1.31E-16 | 1.28E-5 | 3.94E-5 | 1.13E-05 | 3.46E-5 |
| Maximum | 1.25E-15 | 1.02E-14 | 6.29E-4 | 3.44E-3 | 4.52E-4 | 2.22E-03 |

**ANALYSIS S1**

This additional analysis was performed to investigate the risk mitigation effects of an “improved” PCR test with improved diagnostic and/or analytical sensitivity. Improved diagnostic sensitivity refers to an increase in *Se*, in %, and improved analytical sensitivity to a decrease in the limit of detection (*LoD*), in logTCID_50_/aliquot. The simulations were performed using the raw milk farmstore purchasing pathway with methodology identical to that in the main text. Parameter *Se* was evaluated at levels 98.4 (baseline) and 99.9%, and *LoD* at 1.5 (baseline) and 1.0 logTCID_50_/aliquot.

Descriptive statistics for p(infection) are given in **Table S6** and violin plots in **Figure S2**. At both investigated *Se* values, a decrease in *LoD* is shown to have a negligible impact on p(infection). An increase in *Se* is associated with a 0-1-log decrease in infection risk.

The investigated improvement (i.e. decrease from 1.5 to 1.0 logTCID_50_/aliquot) in *LoD* has only slight impact on the risk mitigation effect of PCR testing on the probability of infection from raw milk obtained via farmstore purchase. By contrast, an assumed increase in *Se* (from 98.4% to 99.9%) produces a larger decreases in risk by reducing the probability of an infected bulk tank going undetected (i.e. a false negative). Based on the investigated magnitudes of improvement in the diagnostic and analytical sensitivity, and assuming comparable technical and biological challenges to achieve such improvements, this analysis suggests that benchtop research into molecular diagnostics of H5N1 in milk should prioritize improved diagnostic sensitivity over analytical.

**TABLE S6 | Summary statistics for the scenario analysis of the impact of improved PCR testing parameters limit of detection (*LoD*) and sensitivity (*Se*) on the probability of H5N1 infection per raw milk serving (p(infection)) obtained via farmstore purchase. The values here assume a baseline infected cow diversion (*D*) of 25%.**

|  | ***LoD* 1.5 *Se* 98.4% (Baseline)** | ***LoD* 1.5 *Se* 99.9%** | ***LoD* 1.0 *Se* 98.4%** | ***LoD* 1.0 *Se* 99.9%** |
| --- | --- | --- | --- | --- |
| **Minimum** | 1.35E-17 | 8.44E-19 | 1.35E-17 | 8.44E-19 |
| **5 %ile** | 1.06E-11 | 6.61E-13 | 1.06E-11 | 6.61E-13 |
| **Median** | 2.49E-9 | 1.55E-10 | 2.49E-9 | 1.55E-10 |
| **Mean** | 4.78E-8 | 2.99E-9 | 4.78E-8 | 2.99E-9 |
| **Interquartile Range** | 3.19E-10 - 1.74E-8 | 1.99E-11 - 1.09E-9 | 3.19E-10 - 1.74E-8 | 1.99E-11 - 1.09E-9 |
| **95 %ile** | 2.05E-7 | 1.28E-8 | 2.05E-7 | 1.28E-8 |
| **Maximum** | 1.01E-5 | 6.29E-7 | 1.015E-5 | 6.29E-7 |

**FIGURE S2 | Violin plots for the scenario analysis of the impact of improved PCR testing parameters limit of detection (*LoD*) and sensitivity (*Se*) on the probability of H5N1 infection per raw milk serving (p(infection) obtained via farmstore purchase).** The values here assume a baseline infected cow diversion (*D*) of 25%. For the purposes of visualization, a logarithmic (base-10) Y-axis is employed. The mean is denoted with the red dot.

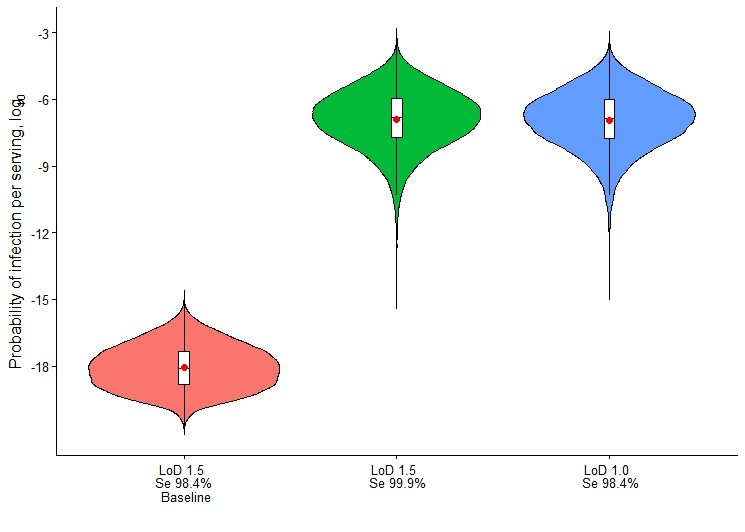

**FIGURE S3 | Diagram of the approach to validation of the pasteurized milk model utilizing the empirical data reported in Spackman et al. (2024b).**

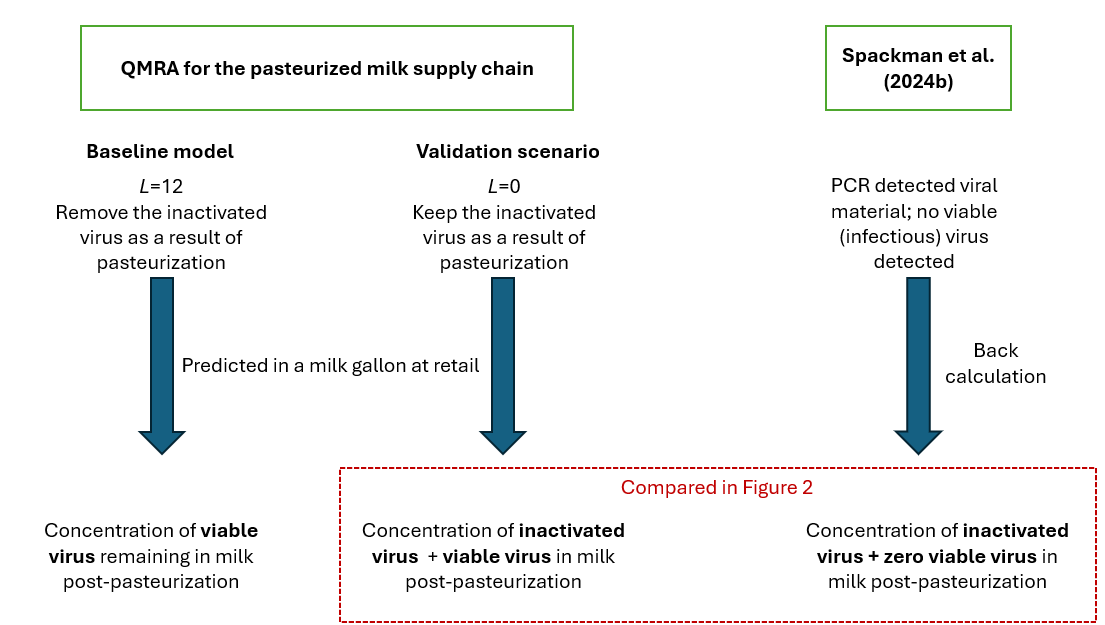

**ANALYSIS S2**

This additional analysis was performed to investigate the numerical difference in logTCID_50_/mL and logEID_50_/mL titrations from the same sample of contaminated milk prior to heat treatment. These assays were performed by coauthors MN and DD. A total of n=6 H5N1-spiked raw milk samples were subject to virus titration by cell culture *in vitro* (logTCID_50_/mL) and embryonated chicken egg (ECE) assays (logEID_50_/mL). The differences between the two values were subject to 2-tailed paired t-test at α=0.05 under the null hypothesis H_0_: (logEID_50_/mL - logTCID_50_/mL) = 0.

**TABLE S7 | Paired viral titrations by cell culture (logTCID_50_/mL) and ECE (logEID_50_/mL) performed on n=6 H5N1-spiked raw milk samples prior to heat treatment.** The resultant p-value was 0.15; as such, we fail to reject the null hypothesis that the numeric difference between these two metrics is 0.

| **LogTCID_50_/mL** | **LogEID_50_/mL** |
| --- | --- |
| 4.72 | 6.11 |
| 4.80 | 5.30 |
| 5.22 | 6.11 |
| 5.14 | 4.97 |
| 4.97 | 4.92 |
| 4.72 | 4.72 |

The above analysis supports the use of logTCID_50_/mL to approximate logEID_50_/mL. Our use of the logEID_50_/mL concentration of viral material in retail fluid milk products reported by Spackman et al. (2024b) in validation of our model is substantiated.

**ANALYSIS S3**

USDA-APHIS does not release the herd sizes of H5N1-infected herds, as this is sensitive and identifying information. To ensure our herd size distribution parameter *H_w_* included the size of larger raw milk dairies, such as Raw Farm, LLC, which experienced a recall due to H5N1 detection in their product, we conducted a supplementary analysis by manipulating *H_W_*. In the baseline raw milk model, *H_W_* is modeled with a PERT(2, 25, 80), based on expert opinion. However, to account for the detection of H5N1 in one of the largest raw milk dairies in the US, we increased the upper limit of the distribution to 1,000 cows. To avoid oversampling of the skewed tail, as the authors and consulted experts agree that the majority of raw milk herds are <100 cows, the distribution was changed to a 4-parameter PERT. This is done in @RISK with the use of the BetaSubj(min, mode, μ, max) function, where μ = (min+max+(λ*mode))/(λ+2) and λ **=** 6 as a skewed shape parameter. The details are available on the Lumivero Website at https://community.lumivero.com/s/article/fourparameter-pert-distribution?language=en_US. The final modified *H_W_* distribution was BetaSubj(2, 25, 144, 1000).

**TABLE S8 | Summary statistics for the scenario analysis of the impact of maximum herd size *H_W_* on the probability of H5N1 infection per raw milk serving (p(infection)).**

|  | **Baseline Maximum Herd Size** | | **Larger Maximum Herd Size** | |
| --- | --- | --- | --- | --- |
|  | Farmstore | Retail | Farmstore | Retail |
| Minimum | 8.50E-16 | 2.22E-15 | 4.83E-16 | 1.34E-15 |
| 5 %ile | 6.67E-10 | 6.65E-10 | 5.44E-10 | 5.33E-10 |
| Median | 1.56E-7 | 1.40E-7 | 1.29E-7 | 1.16E-7 |
| Mean | 2.99E-6 | 2.65E-6 | 2.58E-6 | 2.28E-6 |
| Interquartile Range | 2.00E-8 - 1.09E-6 | 1.82E-8 - 9.45E-7 | 1.63E-8 - 9.03E-7 | 1.48E-8 – 7.84E-7 |
| 95 %ile | 1.28E-5 | 1.13E-05 | 1.09E-5 | 9.49E-6 |
| Maximum | 6.29E-4 | 4.52E-4 | 5.83E-4 | 3.95E-4 |

Increasing the upper limit of *H_W_* had a largely negligible numerical impact on p(infection) in both purchasing pathways. The changes in infection risk per serving is likely driven by the competing mechanisms of dilution, from greater numbers of healthy cows in large herds with low prevalence, versus the extremely concentrated shedding capabilities of infected herdmates. It appears that, in accounting for infrequent large raw milk operations in the population, the effect of dilution exceeds that of shedding in the case of a herd infection. This must also be interpreted in the context of other factors that may be speculated to impact risk, such as: increased regulatory oversight of raw milk farms (higher probability of outbreak detection 🡪 lower risk), and the effect of herd size on probability of outbreak detection (smaller herd 🡪 more individual animal attention 🡪 higher probability of outbreak detection 🡪 lower risk; or, larger herd 🡪 more likely to utilize automated herd health monitoring technologies 🡪 higher probability of outbreak detection 🡪 lower risk).  **ANALYSIS S4**

To determine the relative impact of PCR testing in relation to differing levels of cow diversion (*D*) on the probability of infection per serving (p(infection)) of raw milk obtained through farmstore purchase, an additional simulation was run after setting *%D* at 99.9%, in the absence of PCR testing. The simulation with 99.9% diversion represents a subjectively unrealistic upper maximum meant for comparison of its numerical reduction in risk against that of bulk tank PCR testing.

**TABLE S9 | Summary statistics for the probability of H5N1 infection per raw milk serving (p(infection)) obtained through farmstore purchase in the supplementary analysis of infected cow diversion (parameter *D*) at differing levels.**

|  | **No PCR Testing** | | | | **PCR Testing** |
| --- | --- | --- | --- | --- | --- |
|  | **25% Diversion** | **50% Diversion** | **75% Diversion** | **99.9% Diversion** | **25% Diversion** |
| Minimum | 8.50E-16 | 4.35E-16 | 3.72E-16 | 3.72E-16 | 1.35E-17 |
| 5 %ile | 6.67E-10 | 4.49E-10 | 2.91E-10 | 2.12E-10 | 1.06E-11 |
| Median | 1.56E-7 | 1.06E-7 | 6.98E-8 | 5.06E-8 | 2.49E-9 |
| Mean | 2.99E-6 | 2.08E-6 | 1.40E-6 | 1.07E-6 | 4.78E-8 |
| Interquartile Range | 2.00E-8 - 1.09E-6 | 1.35E-8 - 7.39E-7 | 8.91E-9 - 4.90E-7 | 6.41E-9- 3.55E-7 | 3.19E-10 - 1.74E-8 |
| 95 %ile | 1.28E-5 | 8.83E-6 | 5.89E-6 | 4.37E-6 | 2.05E-7 |
| Maximum | 6.29E-4 | 4.34E-4 | 4.14E-4 | 4.14E-4 | 1.01E-5 |

**FIGURE S4 | Violin plots for probability of H5N1 infection per raw milk serving (p(infection)) obtained through farmstore purchase in the supplementary analysis of infected cow diversion (parameter *D*) at differing levels.** For the purposes of visualization, a logarithmic (base-10) Y-axis is employed. The mean is denoted with the red dot.

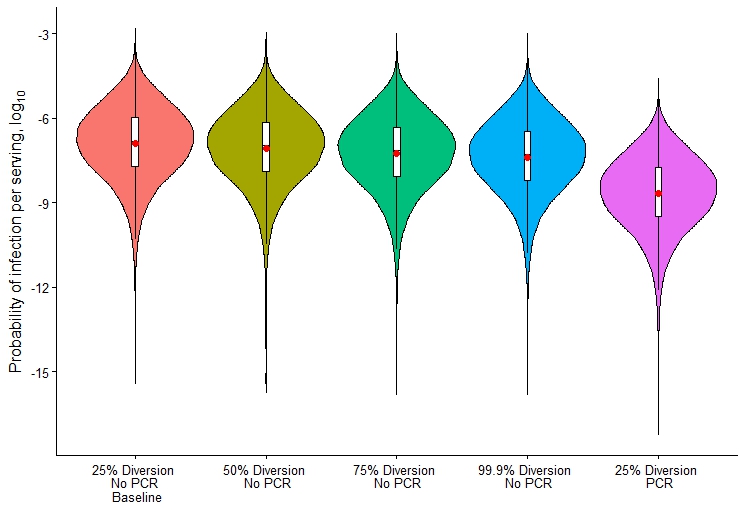

The risk-reducing effect of diversion on p(infection) largely plateaus between 75% and 99.9%. While the distribution of p(infection) shifts downward, the minimum and maximum p(infection) remain unchanged. The minimum, mean, percentiles, median, and maximum of the 25% diversion + PCR testing simulation are lower than those of the 99.9% diversion without PCR testing simulation, although the violin plots in **Figure S4** demonstrate some overlap in values. The interpretation is that further improvement in individual-cow infection detection and diversion capabilities in raw milk herds remain overall less effective than implementation of daily bulk tank PCR testing.

**ASSUMPTIONS**

1. H5N1 clade 2.3.4.4b virions present in the milk of infected cows can infect humans according to an exponential dose-response model such that *p_inf_* = 1-*e*^-Qserv*r^.
2. Clustered distribution of infected herds.
3. Uniform spatial distribution of viruses in fluid suspension.
4. Probability of herd infection does not differ between raw versus conventional herds.

Assumption 1 was evaluated in the uncertainty analysis of parameter *r*. Assumptions 2-4 were evaluated in **Analyses A1-3**, respectively.

**ANALYSIS A1 | Distribution of infected herds**

In the clustered approach to parameter *K* (the number of affected herds in the US), the value for this parameter in each iteration is set by drawing a value for national herd-level prevalence (*p_nat’l_*) and multiplying it by the number of dairy herds in the US according to the 2022 Census of Agriculture. The parameter *p_nat’l_* was arrived at by fitting a distribution based on the range of probabilities of herd infection (# of new H5N1 herd detections within month in a given state / the number of dairy herds within that state) for each positive state for each complete month of the outbreak, using USDA data obtained November 5, 2024. However, this involves extrapolation of national herd-level prevalence from state-level data, and introduces the assumption that *p_nat’l_* does not fall above or below historical within-state herd-level prevalences. As such, we introduced the alternative “homogeneous” approach. In this approach, *K* is parameterized directly using the same USDA data. We obtained the number of new herds with H5N1 infection identified in the US for each month within the dataset, and fit a PERT distribution to this range of values using @RISK. In the homogeneous approach, each iteration draws an integer from this distribution (PERT(10, 56, and 192)) to use as *K* in the “number of infected herds shipping to plant” hypergeometric distribution (**Eq. 1**). However, this approach assumes that any given infected herd is equally likely to ship milk to the modeled processing facility and that the n=100 herds shipping to the processing plant are chosen independently and at random from the entire population of US dairy herds, which is not realistic due to interstate milk shipping regulations, and the contagious nature of the outbreak. However, this is a necessary simplifying assumption when the geographic origin of milk is unknown, as it is in our model. As such, we chose to utilize the clustered approach in the baseline model, but to investigate the impact of the homogeneous geographic distribution assumption in this supplemental analysis.

**TABLE 2 | Summary statistics for the model outputs calculated with two different methods for parameterizing the number of H5N1-infected herds in the US (*K*).** The clustered approach indirectly accounts for the clustering phenomenon of localized outbreaks amongst herds in a geographic area. The homogeneous approach assumes a uniform geographic distribution of infected herds in the US. *Note: 0.00 logTCID_50_ = 1 TCID_50_ (“log” indicates log_10_ transformation)

|  | **Clustered** (Baseline) | | | | **Homogeneous** | | | |
| --- | --- | --- | --- | --- | --- | --- | --- | --- |
|  | Probability of Infection per Serving  (p(infection))  (unitless) | Quantity of Virus per Contaminated Serving  (Q_serv_) (logTCID_50_) | Probability of Serving Contamination  (p(serving))  (unitless) | Number of H5N1-infected herds in US (K) (herds) | Probability of Infection per Serving  (p(infection))  (unitless) | Quantity of Virus per Contaminated Serving  (Q_serv_) (logTCID_50_) | Probability of Serving Contamination  (p(serving))  (unitless) | Number of H5N1-infected herds in US (K) (herds) |
| Minimum | 1.51E-21 | 0.00* | 1.58E-9 | 19 | 7.17E-22 | 0.00* | 7.54E-10 | 10 |
| 5 %ile | 2.39E-20 | 0.00 | 2.43E-8 | 74 | 5.84E-21 | 0.00 | 5.98E-9 | 24 |
| Median | 7.66E-19 | 0.00 | 7.67E-7 | 724 | 8.51E-20 | 0.00 | 8.56E-8 | 67 |
| Mean | 8.71E-18 | 0.019 | 8.66E-6 | 927 | 6.90E-19 | 0.019 | 6.83E-7 | 71 |
| Interquartile Range | 1.41E-19 - 4.39E-18 | 0.00 - 0.00 | 1.41E-7 - 4.41E-6 | 323 - 1338 | 2.21E-20 - 4.25E-19 | 0.00 - 0.00 | 2.24E-8 - 4.27E-7 | 45 - 93 |
| 95 %ile | 4.02E-17 | 0.30 | 4.03E-5 | 2475 | 3.22E-18 | 0.30 | 3.23E-6 | 130 |
| Maximum | 1.25E-15 | 0.30 | 1.32E-3 | 4924 | 9.66E-17 | 0.30 | 6.01E-05 | 192 |

**ANALYSIS A2 | Clustering of viruses in fluid suspension**

Nauta (2005) gives a comprehensive analysis of the mathematical consequences of clustering of infectious agents in risk assessment. Due to the liquid nature of fluid milk, we assume no clustering (i.e. spatial grouping) of virions within the colloidal suspension. However, this assumption must be further investigated by introduction of a clustering parameter into the Gamma function that dictates infectious particle distribution during partitioning. While the baseline scenarios are without clustering (clustering parameter = 1), in **Table 3** we provide results after setting this parameter at 10, therefore representing slight clustering. Clustering is expected to decrease the prevalence of, but increase the quantity of, infectious agent in contaminated servings. The resultant impact on infection risk may be increased, decreased, or remain unimpacted. In our models, with such small quantities of H5N1 virions in contaminated servings, the impact of arbitrary clustering on p(infection) is negligible.

**ANALYSIS A3 | *p_nat’l_* differs between raw and conventional herds**

By using the same distribution for *p_natl’l_* in both raw milk and conventional herds, we inherently assume that no proclivity for H5N1 infection exists between these two types of operations. Various factors may confound milk marketing and H5N1 risk, including herd size (i.e. smaller sizes of raw milk herds translating to higher probability of detecting an ill cow, or lower probability of infection introduction). While no effect could be introduced into the baseline model without arbitrary parameterization, herein we describe the impact of a theoretical ± change in the probability of herd infection for raw milk herds compared to those contributing to pasteurized milk supply chains. As expected, increases/decreases in the probability of herd infection result in directly proportional changes in p(infection); however, the magnitude of these changes is only 0-1 log and as such, the impact of this assumption on our risk projections was deemed to be negligible.

**ANALYSIS 5**

Nooruzzaman et al. (2025a) conducted H5N1 viral decay assays in triplicate in fluid milk at 4, 20, 30, and 37°C, monitoring the titers via egg inoculation conducted over a maximum of 56 days. The authors provided to us the raw data, from which we calculated the decay rate (logTCID_50_/day) for each of the replicates at each temperature. We plotted these decay rates as a function of temperature and fit an exponential line-of-best-fit (R^2^=0.95) to the scatterplot, which resulted in **Eq. 4**: a model that represents viral decay rate *V_D_* as a function of temperature, so we can dynamically and mechanistically model viral decay during storage/transport steps with stochastic distributions for temperature.
